# Guadecitabine plus ipilimumab in unresectable melanoma: five-year follow-up and correlation with integrated, multiomic analysis in the NIBIT-M4 trial

**DOI:** 10.1101/2023.02.09.23285227

**Authors:** Teresa Maria Rosaria Noviello, Anna Maria Di Giacomo, Francesca Pia Caruso, Alessia Covre, Giovanni Scala, Maria Claudia Costa, Sandra Coral, Wolf H. Fridman, Catherine Sautès-Fridman, Roberta Mortarini, Silvia Brich, Giancarlo Pruneri, Elena Simonetti, Maria Fortunata Lofiego, Davide Bedognetti, Andrea Anichini, Michele Maio, Michele Ceccarelli, the EPigenetic Immune-oncology Consortium AIRC (EPICA) investigators

**Author notes:** Contributed equally (Teresa Maria Rosaria Noviello, Anna Maria Di Giacomo, Michele Maio, Michele Ceccarelli).

## Abstract

Association of DNA hypomethylating agents (DHA) with immune-checkpoint inhibitors (ICI) is a promising strategy to improve efficacy of ICI-based therapy. Here we report the five-year clinical outcome and an integrated multi-omics analysis of pre- and on-treatment lesions from advanced melanoma patients enrolled in the phase Ib NIBIT-M4 study, a dose-escalation trial of the DHA agent guadecitabine combined with ipilimumab. With a minimum follow-up of 45 months the median OS was 25.6 months; the 5-year OS rate was 28.9% and the median DoR was 20.6 months. Specific genomic features and extent of T and B cellmediated immunity discriminated lesions of responding compared to non-responding patients. Enrichment for proliferation and EMT-related gene programs, and immune escape mechanisms characterized lesions from non-responding patients. Integration of a genetic immunoediting index (GIE) with an adaptive immunity signature (ICR) stratified patients/lesions into four distinct subsets and discriminated 5-year OS and PFS. These results suggest that coupling of immunoediting with activation of adaptive immunity is a relevant requisite for achieving long term clinical benefit by epigenetic immunomodulation in advanced melanoma patients.

## Introduction

Treatment with immune checkpoint inhibitors (ICI) has dramatically improved the clinical outcome of patients with tumors of different histotypes^1^, including melanoma^2^ and lung cancer^3^. However, the percentage of subjects who benefit from ICI therapy is still low, and novel therapeutic strategies are eagerly awaited to fully exploit their clinical potential. Indeed, even in the most responsive tumor types, both intrinsic^4^ and acquired resistance^5,6^ limit the efficacy of ICI therapy, and the development of more effective ICI-based treatments is hindered by incomplete knowledge of the genetic mechanism governing host–tumor interaction^7,8^. Nevertheless, the cellular and molecular characterization of human tumor samples by highthroughput and deep phenotyping approaches define the role of the immune microenvironment in driving the prognosis of cancer patients and their responsiveness to ICI therapies^9,10^.

In this scenario, an active area of biomedical research aims to identify combinatorial approaches that could improve even the early phases of developing the anti-tumor response. Mechanistically, these new immunotherapy regimens aim at achieving one or more of three main effects thought to be crucial for overcoming resistance to immune intervention: fostering the cross-talk between innate and adaptive arms of the immune system, promoting the recruitment of functional T cells at the tumor site, and counteracting recruitment/function of immunosuppressive cells^11^.

Among novel agents that may play a role in ICI combinations are the demethylating agents (DHA) due to their immunomodulatory activity on tumor cells^12^, the ability to activate innate immunity pathways^13,14^ and the pre-clinical evidence for enhanced anti-tumor effects when combined with ICI^15^. In this scenario, our Italian Network for Tumor Biotherapy (NIBIT) Foundation Phase Ib NIBIT-M4 trial based on the association of ipilimumab with the DNA hypomethylating agent (DHA) guadecitabine in advanced melanoma patients^13^, showed significant tumor immunomodulatory effects and preliminary evidence of promising clinical activity. More recently, by comparing transcriptional programs elicited by different classes of epigenetic drugs in melanoma cells, we found that the main biological activity of guadecitabine is the promotion of gene expression and activation of master factors belonging to innate immunity pathways, including Type I-III IFN, NF-kB and TLR^12^. These results corroborated the notion that rescue of adaptive immunity by ICI may cooperate with the promotion of innate immunity by the DHA guadecitabine, thus potentially explaining the clinical activity of the combination. Two additional recent trials, combining guadecitabine with pembrolizumab in solid tumors^16,17^ have indeed confirmed the significant anti-tumor activity of this combination in terms of clinical benefit rate (31.4%) or of progression-free survival rate >24 weeks (37%). Crucially, in both studies, relevant immune effects were described regarding the upregulation of innate and adaptive immunity pathways in post-treatment samples.

Despite these initial promising clinical applications of the DHA guadecitabine combined with ICI, the further development of these combinatorial approaches opens the question of understanding the biology of response and resistance to this specific type of epigenetic immunomodulation. To this end, we developed an advanced integrative systems biology approach aiming at characterizing baseline and on-treatment tumor tissues from NIBIT-M4 patients. The results of this multi-omics profiling revealed that long-term clinical benefit might be predicted by a combinatorial index integrating transcriptional information on the development of adaptive immunity with an index of genetic immunoediting.

Previous studies have shown that genetic evidence of neoantigen depletion is preferentially observed in lesions with high Immunoscore and that immunoedited tumor clones do not recur, while progressing clones tend to be immune privileged, despite the presence of tumor-infiltrating lymphocytes^18^. We quantify the amount of genetic immunoediting (GIE) as the ratio between observed and predicted tumor neoantigens. The presence of an adaptive immune response within tumors is accounted for by the Immunological Constant of Rejection (ICR) ^19,20^. This signature predicts survival and response to ICI in different tumors such as Breast^21^, Bladder, Stomach, Head and Neck^20^, Sarcoma^22^, and Melanoma^9^. The stratification induced by the ICR/GIE classification predicts response in our cohort and larger ICI studies. Our data can serve as a guide for improved patient stratification and selection strategies.

## Results

### Long-term outcomes in the NIBIT-M4 trial

At data cut-off, July 1^st^ 2022, with a minimum follow-up of 45 months, 6 (31%) of the 19 patients enrolled in the NIBIT-M4 study were alive. The median OS was 25.6 months (95% CI, 0.0-52.9), while the median PFS was 5.2 months (95% CI, 4.0-6.4); the 5-year OS rate was 28.9% with a 5-year PFS rate of 5.3%; median DoR was 20.6 months (95% CI, 12.4-28.8). Three patients were in CR and off study therapy, while 13/19 (68%) had received subsequent line(s) of therapy; among those, 5 patients who had achieved a DCR had a median time to subsequent treatment of 18.9 months (range 10.3-39.0) (**Figure 1**).

**Figure 1.**
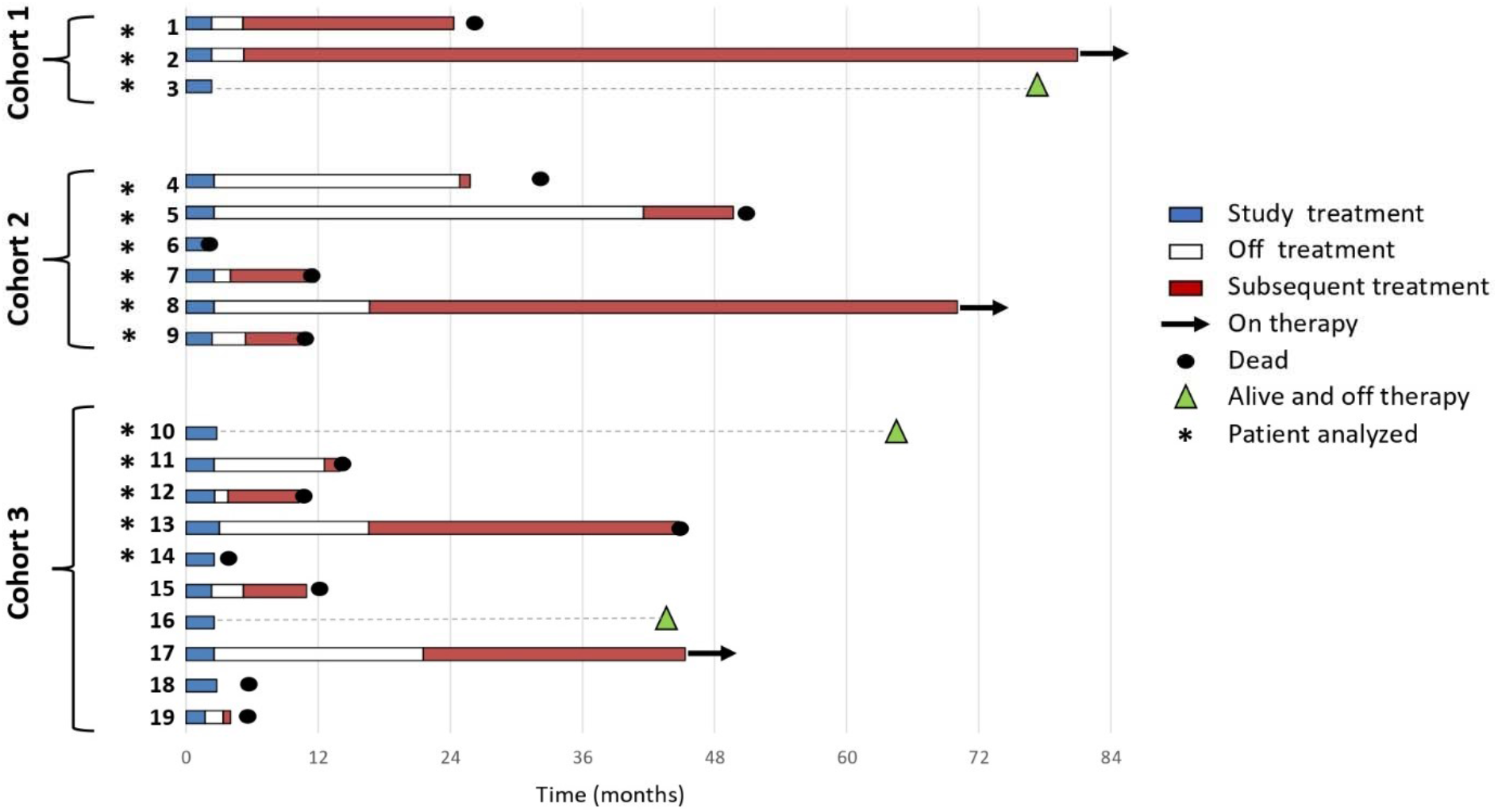
Swimmer plot analysis of NIBIT-M4 patients. Swimmer plot showing by study Cohort patients who at the time of data cut-off were alive and off study therapy without having received subsequent treatment (n=3), patient who died without receiving subsequent treatment (n=3), and all patients who had received subsequent treatment at the time of data cut-off, regardless of whether alive or dead (n=13). Subsequent treatments included: anti-PD1 as monotherapy or in combination, BRAFi+MEKi, ICOS agonist, chemotherapy.

### Genomics landscape: mutational profile differences in baseline and on-treatment lesions in R vs. NR patients

Longitudinal multi-omics profiling, including Whole Exome Sequencing (WES), RNA Sequencing (RNASeq), and Reduced-representation bisulfite sequencing (RRBS), were performed on tumor biopsies collected at baseline (week 0) and week 4 and week 12 on therapy from 14 patients (**Supplementary Figure 1**). Matched normal tissue collected at baseline was available for 8 patients. The exome sequencing profiling of our cohort, performed using stringent filtering, showed a high consistency of the somatic calls/mutations during treatment (**Figure 2, Supplementary Table 1**). Although tumor mutational burden (TMB) was not significantly different in R vs NR patients (16.9 vs. 15.3, *p*-value 0.6), several significant differences were found at the single gene level. *BRAF* was slightly enriched in NR (*p*-value 0.02). In contrast, *NRAS* mutation was significantly more frequent in R vs. NR (50% vs. 0%, *p*-value 5.4e-5). *ADAMDEC1*, encoding a disintegrin metalloproteinase associated with dendritic cell function, was altered only in lesions from R patients. Mutations in genes belonging to the epithelial to mesenchymal transition (EMT) pathway were enriched in NR (*p*-value 0.01), in agreement with EMT being associated with a less favorable outcome in patients with cancer^23^. Three neuronal-related genes (*PCLO, PLXNA4*, and *EPHA7*), and the gene encoding the leptin receptor (*LEPR*), all reported as mutated in melanoma at a variable frequency (37%, 11%, 16% and 8% of samples in TCGA cohort, respectively), were altered more frequently in R compared to NR patients. The *CDKN2A* gene, frequently mutated in melanoma (35% of patients in TCGA cohort), was more frequently altered in NR patients. The *DNMT1* gene, encoding one of the guadecitabine targets, was mutated in two NR patients, and one of the mutations was a truncating event suggesting loss of function of the *DNMT1* gene product. Finally, the male germline-specific gene *PLCZ1* was mutated only in lesions from R patients.

**Figure 2.**
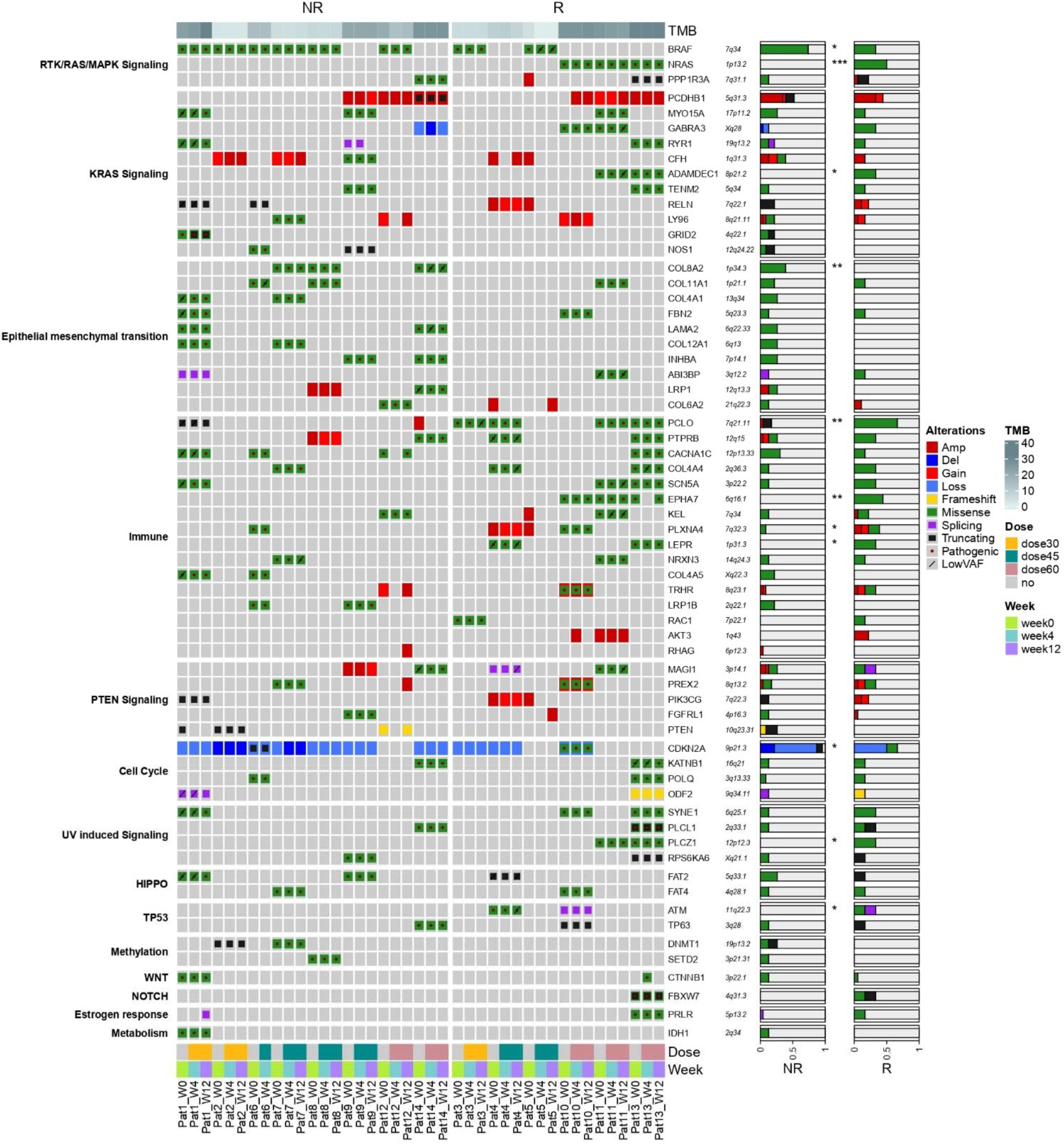
Genomics Landscape of the NIBIT-M4 trial. Oncoplot of frequent somatic nonsynonymous and copy number alterations of NIBIT-M4 trial organized by response (columns) and pathways (rows). Tumor Mutation Burden (TMB), dose (30, 45 and 60 mg/m2/day) and time (week0, week4, week12) of treatments are indicated. Proportion of alterations in non-responder (NR) and responder (R) groups is visualized for each gene (p-value of Pearson’s chi-squared test statistic, *: p <0.05, **: p < 0.01, ***: p < 0.001).

### Transcriptional landscape of baseline and on-treatment tumor lesions: distinct and evolving transcriptional programs distinguish R from NR patients

RNA-sequencing data from the NIBIT-M4 were used to carry out differential gene expression analysis between R and NR patients at different time points of treatment (**Supplementary Table 2**). This analysis showed a progressive enrichment from baseline to week 12 in Gene Ontology Biological Processes (GO:BP) categories related to immune processes in R compared to NR patients (**Figure 3A** and **Supplementary Table 3**).

**Figure 3.**
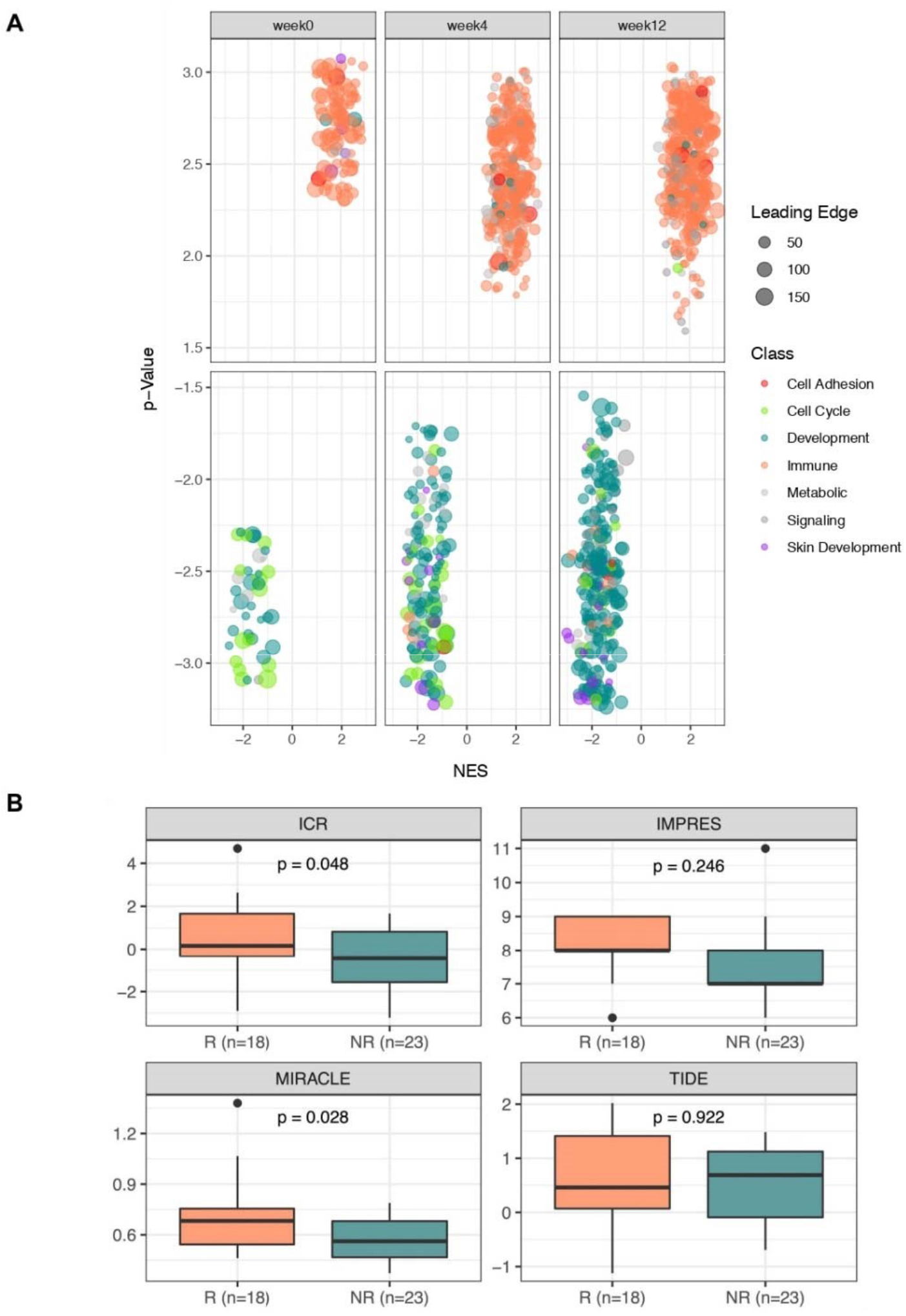
Transcriptional Landscape of the NIBIT-M4 trial (1) Supervised differential analysis between responders and non-responders before treatment (week0) and after four (week4) and twelve (week12) weeks. Enriched GO terms are visualized as dotplot, grouped in categories and divided between responders (top) and non-responders (bottom). Axes show the Normalized Enrichment Score (NES) and -log10 of p-value (-log10(p-value)) from GSEA, respectively. Dot size is proportional to the size of the leading-edge subset, i.e. the gene core that accounts for the gene set enrichment signal (A). ICI response prediction scores between responder and non-responder samples (pvalue of Student’s t-Test) (B).

In contrast, in lesions from NR patients, a progressive increase from baseline to week 12 was found for GO terms related to adhesion, cell cycle, metabolism, and skin developmental processes. By testing several state-of-the-art predictive signatures of response to ICI, we found that the MIRACLE (Mediators of Immune Response Against Cancer in soLid microEnvironments) score^9^ and ICR (Immunologic Constant of Rejection)^24^ signature reached statistical significance (*p*-value < 0.05, **Figure 3B** and **Supplementary Table 4**), but not the IMPRES^25^ and TIDE^26,27^. None of these four scores discriminated against R from NR patients when considering only baseline samples (**Supplementary Figure 2A**).

We analyzed the differential expression between R and NR patients in selected gene sets (**Figure 4A**). In agreement with the GO analysis, lesions from R patients showed a progressive increase of expression, from baseline to week12, of genes encoding for molecules controlling T cell activation, inhibitory receptors and ligands, chemokines, HLA class II antigens and components of the immunoproteasome. Several of the genes in the ICR signature^20,24^ showed a progressive expression increase in R patients during therapy, compared to NR patients.

**Figure 4.**
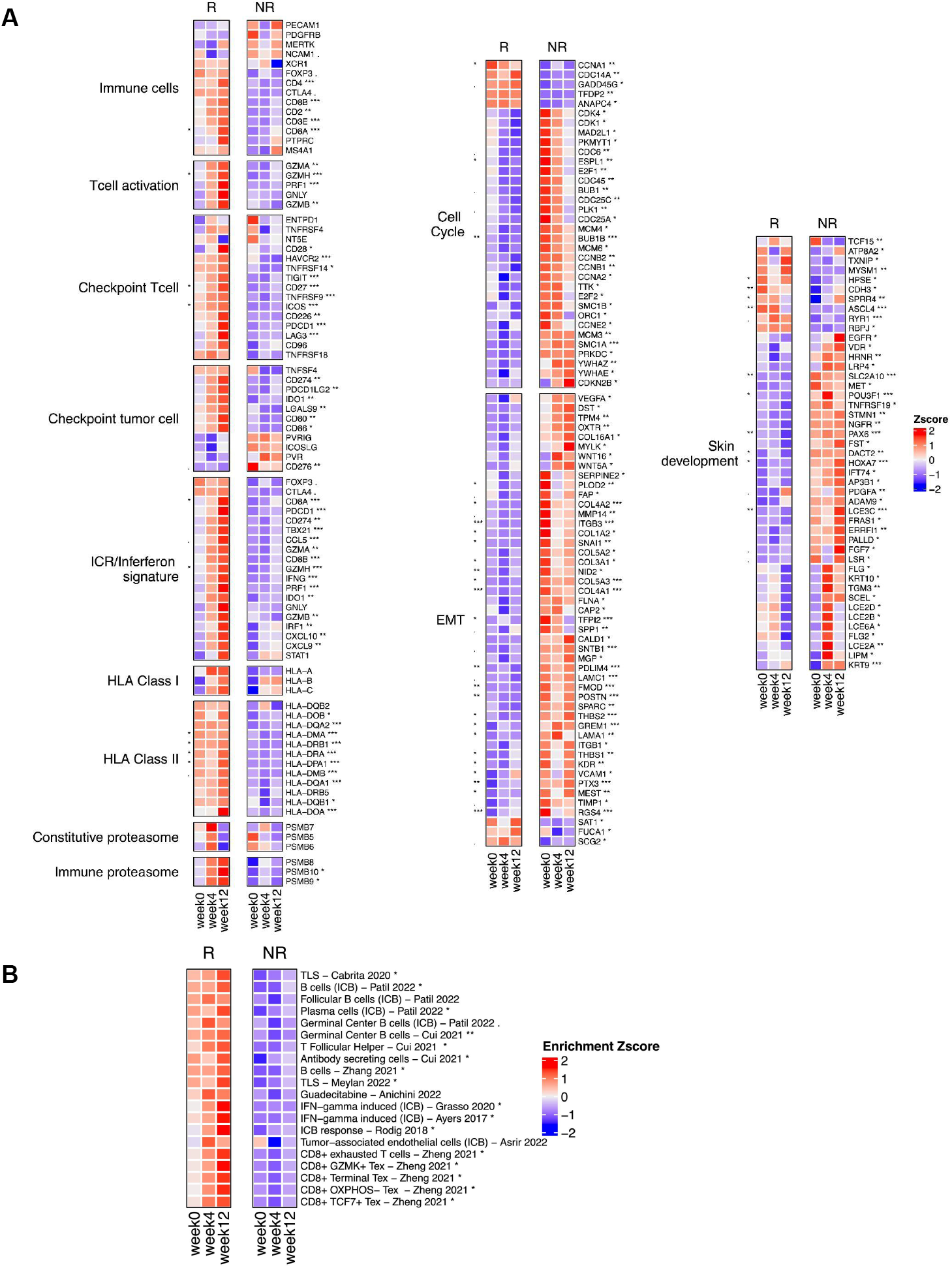
Transcriptional Landscape of NIBIT-M4 trial (2). Heatmap of expression z-scores (from RNASeq profiling) computed for genes belonging to selected pathways between responder and non-responders grouped for treatment timepoints. Significance values on the left correspond to the difference at the baseline (week 0), whereas significance levels on the right of the heatmap correspond to all weeks together (p-value from edgeR differential expression analysis, ·: p <0.1, *: p <0.05, **: p < 0.01, ***: p < 0.001) (A). Heatmap of enrichment z-scores (from NANOSTRING gene expression assay) computed for selected pathways between responder and non-responders grouped for treatment timepoints. Significance values on the right correspond to the difference correspond to all weeks together (p-value from Student’s t-Test, ·: p <0.1, *: p <0.05, **: p < 0.01) (B).

By contrast, lesions from NR patients showed significantly higher expression of cell cycle-, EMT- and skin development-related genes. Several of the EMT-related genes with higher expression in NR patients encoded for molecules controlling adhesion (*ITGB3, VCAM1*, collagens, and others), interaction with extracellular matrix (*VEGFA, MMP14, WNT5A, LAMA1*, and others), and melanoma de-differentiation (*KRT9, KRT10, EGFR*, and others). Lesions from NR patients also showed significantly higher expression, compared to R patients, in several key genes controlling cell proliferation including cyclin-dependent kinases 1 and 4, cyclins B1 and B2, mitotic checkpoint serine/threonine kinase B, and other transcription factors controlling cell cycle such as *E2F1* and *E2F2*.

By a custom-designed NanoString assay we then explored differential expression in R vs NR lesions of 20 published immune-related signatures (**Figure 4B**) providing information on B cell content and differentiation^28,29^, tertiary lymphoid structures (TLS) formation^30,31^, follicular T helper cells^28,29^, T-cell exhaustion (TEX) subsets^32^, tumor-associated endothelial cells^33^, ICB response^34,35^, and the recently identified guadecitabine-specific signature genes induced by this demethylating agent in melanoma cell lines^12^.

The large majority of these signatures were selectively enriched, considering all time points, in tumor biopsies from R compared to NR patients. These results were consistent with preferential development in R lesions of a coordinated T- and B-cell mediated immune response involving TLS and TFH cells, with enhanced expression of IFN-γ-induced genes crucial for ICB response and with increased presence of CD8^+^ T cells at different stages of exhaustion. In agreement with this interpretation, unsupervised clustering of all lesions for the level of expression of these 20 signatures showed that all lesions belonging to the “20signature high profile” clustered together with the “High-ICR” profile (**Supplementary Figure 2B**).

Interestingly, progressive increase in expression of guadecitabine-specific signature genes^12^ in R vs NR patients was observed by both the NanoString assay (**Figure 4B**) and by differential expression analysis at each time point based on RNA-seq data (**Supplementary Figure 2C**).

The composition of the tumor microenvironment of our cohort was deconvolved from transcriptome profiling data using eight immune- and two stromal-cell signatures with MCPcounter^36^ (**Figure 5A**). We found a significantly higher abundance in R vs NR patients of the following immune sub-populations: T cells (*p*-value 0.01), CD8^+^ T cells (*p*-value 0.009), cytotoxic lymphocytes (*p*-value 0.005) and Natural Killer (NK) cells (*p*value 0.04). Interestingly, the specific immune subtypes that discriminated against R from NR lesions also showed a progressive increase over time of treatment in R subjects (**Figure 5A**).

**Figure 5.**
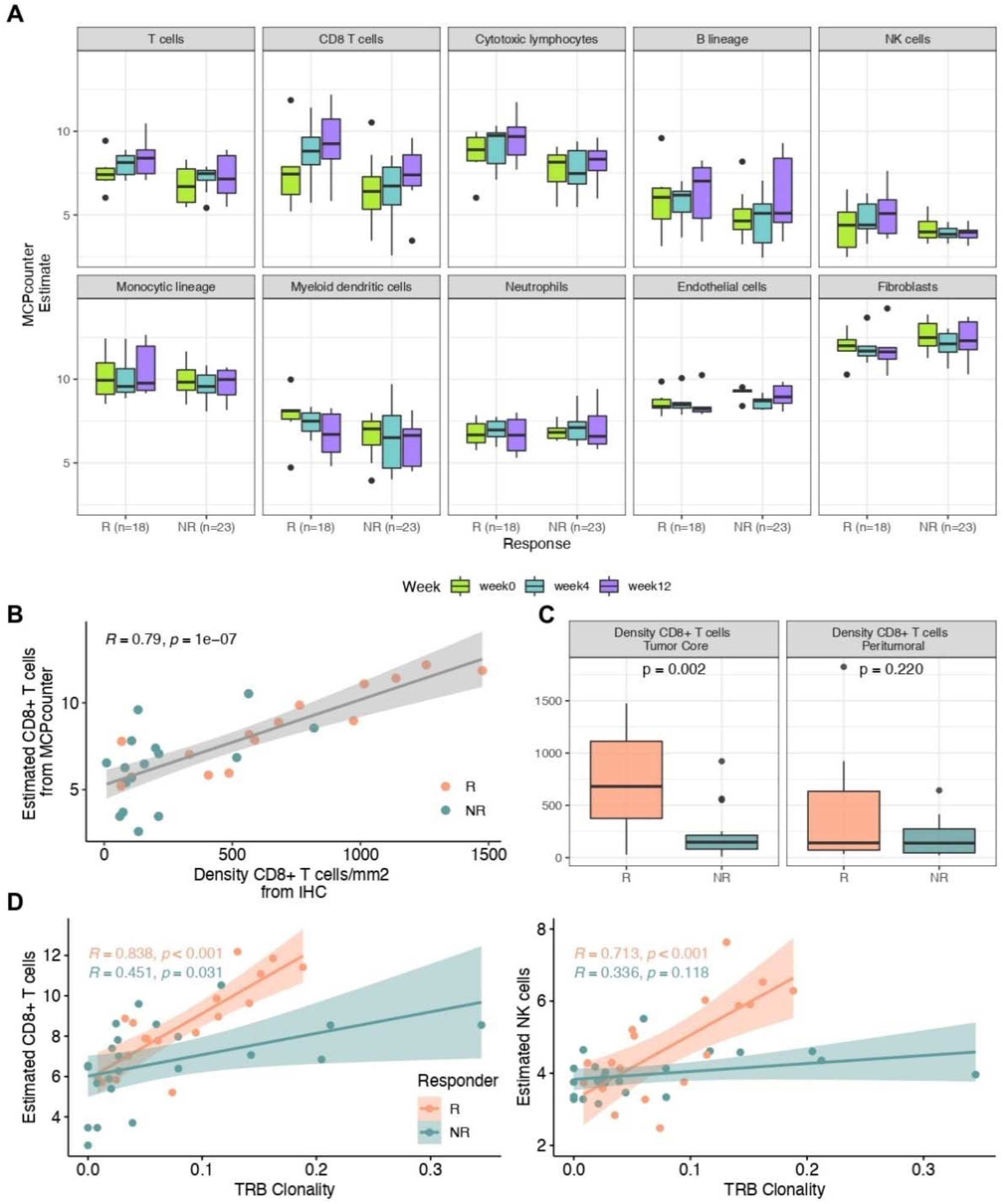
Immune Contexture. Deconvolution of immune cell fractions stratified by time point and response (x axis) (A). IHC validation (x axis) of the deconvolution of CD8 T-cell proportion estimated from RNAseq (y axis) (spearman correlation coefficients rho (R) and associated p-values are provided for responder and non-responder groups) (B). Density of CD8 T-cells by location from IHC (p-value of Student’s t-Test between responder and nonresponder groups) (C). Scatterplot between the T-cell receptor clonality (B locus) and CD8 T-cell abundance (left) and NK cell abundance (right) (spearman correlation coefficients rho (R) and associated p-values are provided for R and NR groups) (D).

The estimated increased abundance of CD8^+^ T cells in R lesions was in agreement with available quantitative immunohistochemistry (IHC) data (R = 0.79, *p*-value 1e-07, N = 11) for this immune subset (**Figure 5B**) and with enhanced CD8^+^ intratumoral T cells in R lesions (*p*-value 0.002) (**Figure 5C**). We then used the clonality of V(D)J rearrangements within the TCR Beta locus (TRB) as a proxy for estimating T cell expansion during therapy. TRB clonality was significantly and highly correlated with the estimated abundances of CD8^+^ T cells (R = 0.838, *p*-value < 0.001) and NK (R = 0.713, *p*-value < 0.001) cells in R rather than NR patients (**Figure 5D**).

Taken together, the gene expression landscape of NIBIT-M4 lesions indicated that distinct and evolving transcriptional profiles characterized baseline and on-treatment tumor biopsies from R compared to NR patients. Lesions from R patients showed progressive enrichment for signatures and gene sets revealing activation of adaptive immunity and effective immunomodulation by guadecitabine with a preferential and clonal activation of T cell subpopulation and NK cell in the tumor microenvironment. Lesions from NR patients revealed lack/defective promotion of immunity in a tumor transcriptional background dominated by proliferation and EMT processes.

### Integrative analysis of methylation and transcriptomic profiles during treatment

A rank aggregation analysis of gene expression and gene methylation in NIBIT-M4 tumor biopsies demonstrated the interdependent changes between gene body methylation levels and gene expression induced by guadecitabine. This analysis for epigenetically regulated pathways confirmed the remarkable promotion of T cell mediated and humoral immunity in lesions from R patients, but not in NR patients, during treatment and enrichment for cell cycle-related and development pathways in NR patients (**Figure 6A**). By exploiting further the expected inverse relationship between gene expression and promoter methylation, we then identified modules of hypomethylated, immune-related genes up-regulated during treatment in R vs NR patients (shown in the upper right quadrant in each plot of **Figure 6B**). Interestingly the number of these genes increased during treatment, consistent with the expected mechanism of action of guadecitabine.

**Figure 6.**
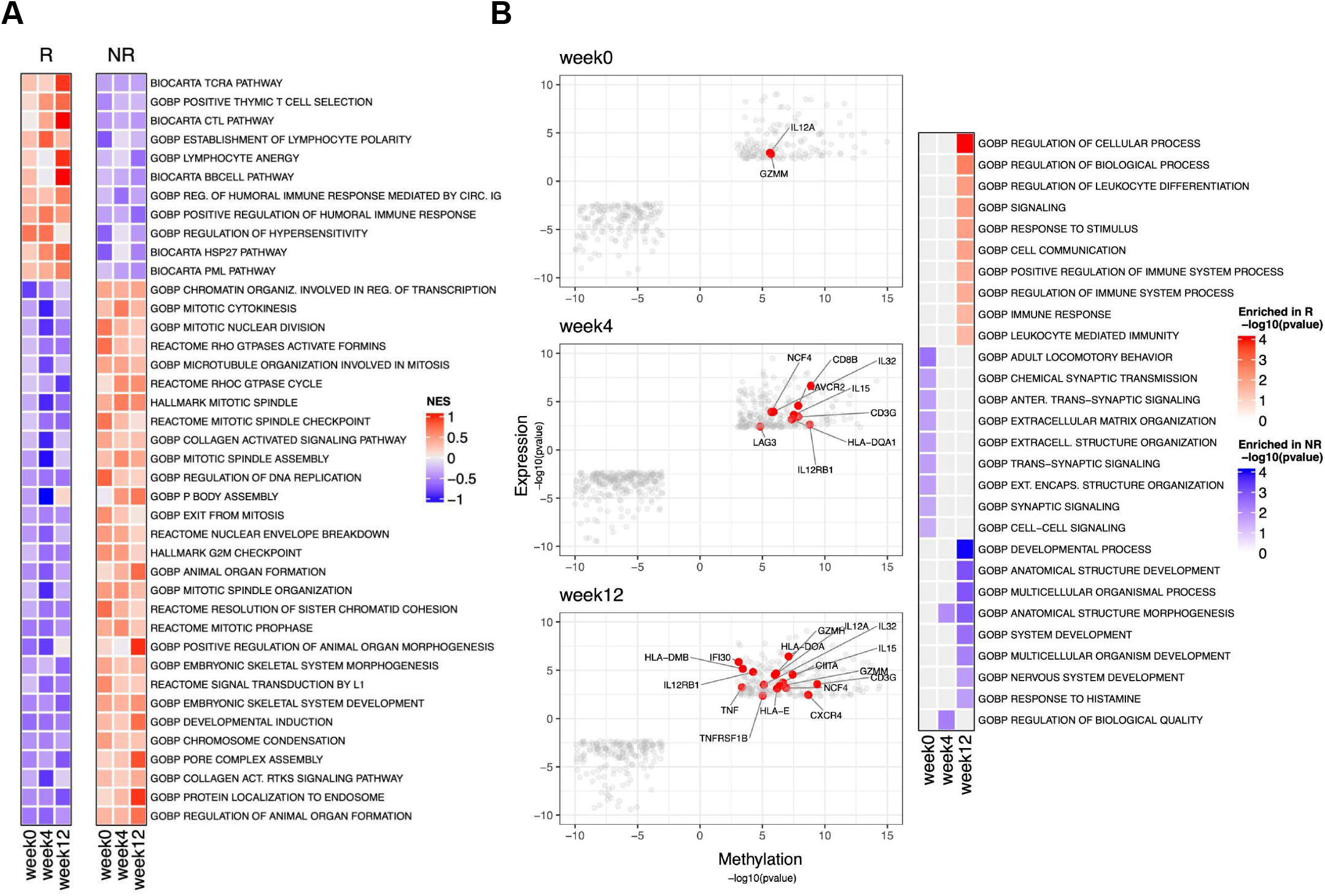
Epigenetic regulation of functional pathways. Normalized Enrichment Score (NES) of epigenetically regulated pathways differentiating responders and non-responders obtained by the integration of the gene expression and gene methylation ranks (A). Starburst plot of promoter hyper-methylated and down-regulated genes (lower-left quadrant), and promoter hypo-methylated and up-regulated genes (upper-right quadrant) between responders and non-responders at different weeks of treatment (left). Heatmap of enrichment scores as -log10(p-value) from GO:BP Overrepresentation analysis for hypo-methylated and up-regulated genes between responders and nonresponders at different weeks of treatment (right) (B).

### The ICR/GIE classification contributes to explain response, resistance through immune escape and long term clinical outcome

TMB and estimated neoantigen loads were highly correlated (R=0.77, *p*-value 5e-09) in the lesions of the NIBIT-M4 trial, however none of these two parameters discriminated against R from NR patients.

We then tested the hypothesis that a combined index, the genetic immunoediting (GIE) score^37,38^, could instead show an association with clinical response. GIE index integrates both TMB and neoantigen load information in a single measure of the extent of immunoediting.

The GIE value was calculated as the ratio between the observed vs. expected number of neoantigens in each tumor sample (**Supplementary Table 5**) The expected number of neoantigens was estimated by training a linear model having TMB as the independent variable and neoantigen load as the outcome variable. Tumors with a number of neoantigens lower than expected (i.e., lower GIE values) are thought to display evidence of immunoediting, whereas a higher frequency of neoantigens than expected indicates a lack of immunoediting (Non-GIE).

In agreement with our hypothesis, when taking into account all available lesions, R patients had a significantly lower GIE score than NR patients.

However, the difference was not significant at the baseline, possibly due to the low number of cases (**Supplementary Figure 3A**). We then correlated the ICR signature score ^19,20^ with GIE index values, but no clear correlation was observed. We then stratified tumor lesions from patients enrolled in the NIBIT-M4 study based on GIE score greater or lower than one and ICR score greater or lower than zero, yielding four groups: High-ICR/GIE, High-ICR/Non-GIE, Low-ICR/GIE, Low-ICR/Non-GIE (**Figure 7A**). The tumor samples belonging to R patients were highly enriched in the High-ICR/GIE group (61%, *p*-value 1.4e-04). The ICR/GIE classification was significant even when limiting the analysis on baseline lesions (67%, *p*-value 2.1e-02) with four of the five lesions in the High-ICR/GIE group (**Supplementary Figure 3B**). To shed light on the mechanism that differentiates GIE in the presence of adaptive immunity captured by ICR, we then performed a supervised transcriptome analysis comparing the High-ICR/GIE vs. the High-ICR/Non-GIE groups (**Figure 7B**). This analysis showed that the group High-ICR/GIE was characterized by enhanced representation of several GO:BP categories related to immune response including antigen processing and differentiation. This suggested to us that the “High-ICR/Non-GIE” could be defective for expression of antigen processing and presentation genes in a way that could explain the lack of immunoediting. In agreement, lesions from the High-ICR/Non-GIE group showed loss of expression of HLA class I antigens on tumor cells despite the presence of CD8^+^ T cells infiltrating the tumor core (**Figure 8** and **Supplementary Figure 4A**). Comprehensive assessment of all available lesions from 11 patients for the ICR, GIE, CD8^+^ and HLA Class I scores (**Supplementary Figure 5A**) indicated that the overall profile for these four parameters was consistent with the observed clinical response in all but one patient (#11). Patient #11, had a low ICR/non GIE score, but was classified in the responder subset due to stable disease in target lesions. Eventually the patient underwent disease progression in target lesions and developed several new lesions (**Supplementary Figure S5B**). Collectively, these findings provided a mechanistic explanation for the genesis of the High-ICR/Non-GIE subset. In fact, lesions with defective expression of HLA class I molecules may retain evidence for development of adaptive immunity (High-ICR) and also for CD8^+^ T cell infiltration, but downmodulation of MHC class I molecules on tumor cells prevents recognition of HLA/neoantigen complexes by T cells, thus suppressing the possibility of immunoediting (therefore the lesions are identified as Non-GIE).

**Figure 7.**
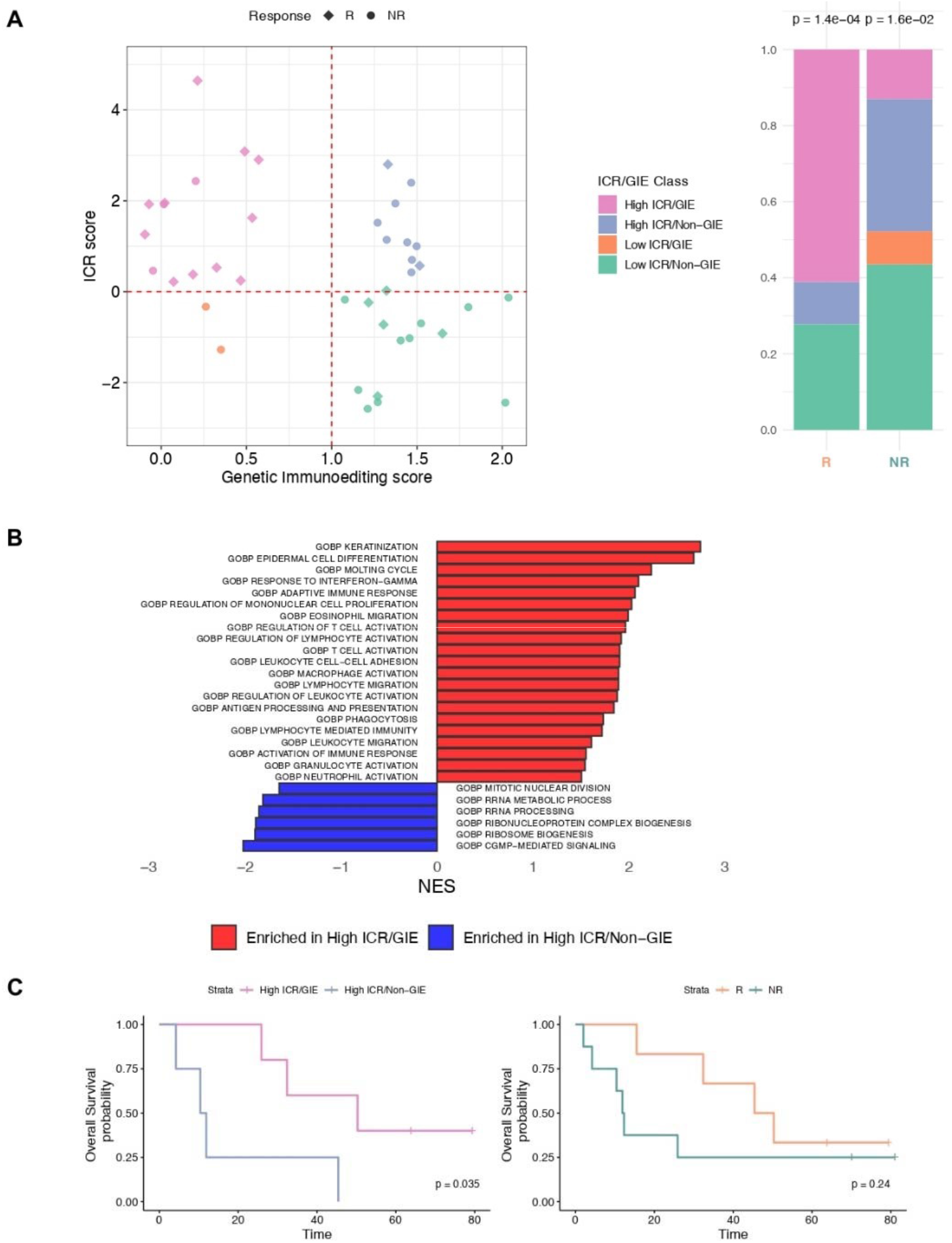
Biomarkers of Immuno-Editing (1). Scatterplot of ICR score by Genetic Immunoediting (GIE) score for responder and non-responder samples (left) and their proportion (right) after classification as ICR/GIE classes in responder and non-responder groups (p-value from Pearson’s chi-squared test statistic) (A). Barplot of most significantly (FDR < 0.01) enriched GO:BP categories from GSEA analysis of High ICR/GIE vs. High ICR/Non-GIE comparison (B). Kaplan Meier for OS by patients classified as High ICR/GIE or High ICR/Non-GIE at week12 (left) and responder or non-responder (right). Time is indicated in months and censor points are indicated by vertical lines (C)

**Figure 8.**
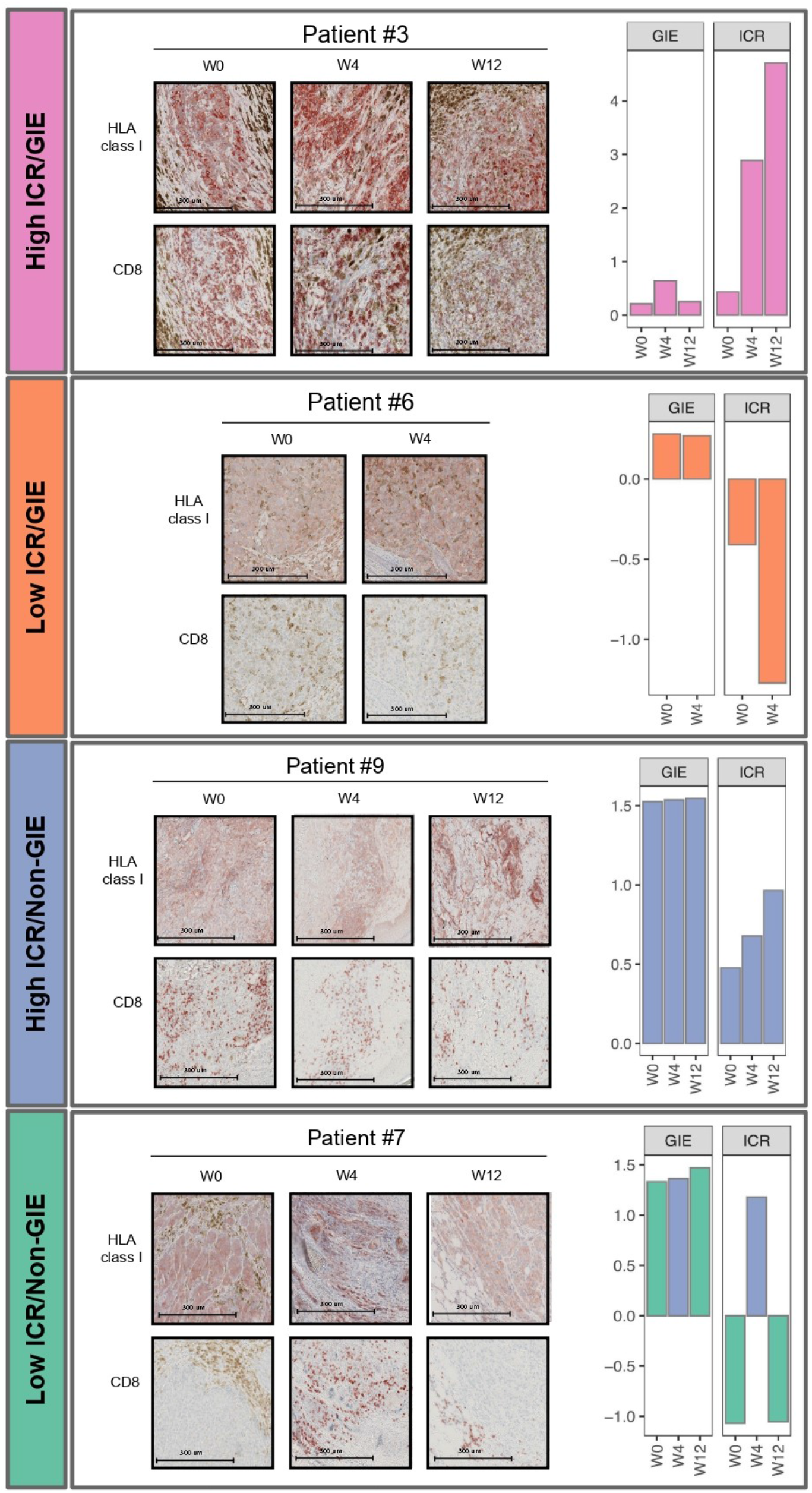
Biomarkers of Immuno-Editing (2). Microphotographs of HLA class I and CD8 immunohistochemistry for representative patients in each ICR/GIE class (left) and ICR and GIE sample scores for each time points (right)

We then asked whether the patients groups defined by the ICR/GIE classification also experienced different long term clinical outcomes. By stratifying patients according to the ICR/GIE classification of week 12 biopsies we found a significant difference (*p*-value 0.035) in overall survival (OS) and in PFS between the High ICR/GIE and the High ICR/Non-GIE group (**Figure 7C** and **Supplementary Figure 4B**, left hand panel). Both OS and PFS were significantly discriminated by the ICR/GIE stratification even when taking into consideration all four subsets. In contrast, patients’ classification by response groups was not associated with OS, although it was associated with PFS (**Figure 7C** and **Supplementary Figure 4B**, right hand panel).

To validate the ICR/GIE stratification, we assembled a cohort of 83 melanoma cases treated with ICI, either anti-CTLA4 or -PD1, from previous published studies^4,39,40^ for which TMB, neoantigen load and gene expression were available. This analysis confirmed the stratification of patients into 4 subsets as seen in the NIBIT-M4 cohort (**Figure 9A)**. Significant survival differences between lesions characterized as HighICR/GIE vs. those coded as High-ICR/non-GIE were observed even in this validation cohort (**Figure 9B**). Interestingly, differential GO:BP pathway analysis in the High-ICR/GIE vs High-ICR/non-GIE subsets identified the GO:BP category “antigen processing and differentiation” as selectively enriched in the HighICR/GIE subsets as we found in the NIBIT-M4 cohort (**Figure 9C**). These results suggest that the same mechanism uncovered in the NIBIT-M4 cohort could explain the High ICR/Non-GIE subset even in the validation cohort: a defective antigen processing and presentation pathway may contribute to suppress immunoediting even when lesions have a high ICR profile, and this may be a general phenomenon irrespective of the type of immunotherapy that is being used.

**Figure 9.**
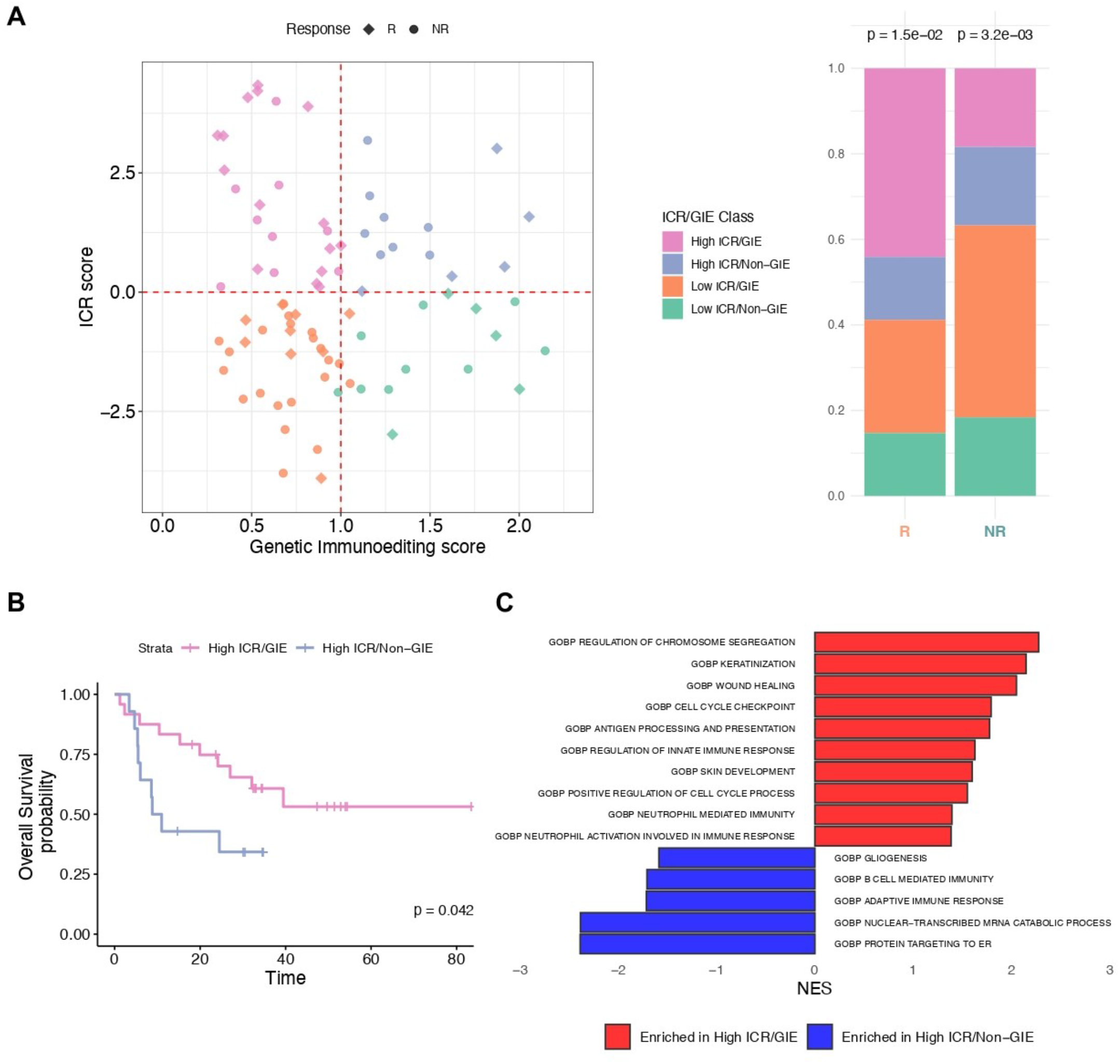
Validation of the ICR/GIE score in three independent cohorts. Scatterplot of ICR score by Genetic Immunoediting (GIE) score for responder and non-responder samples (N=83) (left) and their proportion (right) after classification as ICR/GIE classes in responder and nonresponder groups (p-value from Pearson’s chi-squared test statistic) (A). Kaplan Meier for OS by patients classified as High ICR/GIE or High ICR/Non-GIE. Time is indicated in months and censor points are indicated by vertical lines (B). Barplot of most significantly (FDR < 0.01) enriched GO:BP categories from GSEA analysis of High ICR/GIE vs. High ICR/Non-GIE comparison (C).

Collectively, these results suggest that effective coupling of tumor immunoediting with activation of adaptive immunity can promote response and improved clinical outcome in the NIBIT-M4 epigenetic immunomodulation trial. In contrast, defective development of adaptive immunity or lack of immunoediting, associated with immune escape mechanisms, may favor resistance and less favorable long term clinical outcome.

## Discussion

The NIBIT-M4 trial has been the first Phase Ib epigenetic immunomodulation study testing the association of the DHA guadecitabine with ICI in solid malignancies. Treatment of metastatic melanoma patients with guadecitabine combined with ipilimumab was found to be safe, feasible and tolerable, with initial signs of clinical and immunologic activity^13^. The 5-year survival rate, duration of response, and time to subsequent treatment in patients who achieved a disease control we report here are intriguing and clinically meaningful. These findings compare favorably with the efficacy of CTLA-4 monotherapy in metastatic melanoma patients^41^, though with the limitations of interstudy comparisons that need to be interpreted with caution and to be placed in context. Nevertheless, the long-term follow-up of the NIBIT-M4 trial, seems to support the clinical potential of guadecitabine combined with CTLA-4 therapy, though the relative contribution of each agent and the potential of guadecitabine maintenance therapy in the clinical results we observed could not be fully dissected in this trial. In this scenario, our ongoing randomized phase II NIBIT-ML1 study (NCT04250246) in PD-1 resistant melanoma patients will help to address the clinical and immunobiologic contribution of the addition of the DHA ASTX727 to ICI therapy. However, further support the notion that guadecitabine is a promising way forward to improve the efficacy of ICI therapy in solid tumors derives from two most recently published trials in platinum-resistant ovarian cancer^16^, and in different tumor types^17^. Indeed, the combination of guadecitabine with the anti-PD-1 pembrolizumab led to encouraging response rates, immunomodulation in the tumor tissue and/or in periphery, evidence of demethylation in on-treatment lesions, with manageable toxicity^16-17^.

Our aim here was to exploit the complete longitudinal multi-omics profiling of the whole NIBIT-M4 cohort in conjunction with the five years follow-up to shed light on the effect of the combination during treatment and to identify early biomarkers of response. To this end, we have first individually interrogated the available omics platforms taking advantage of the accurate longitudinal sampling. Then we developed innovative computational multi-omics integration approaches to evaluate the effect of the adopted demethylation agent on boosting the adaptive and innate immune-mediated cancer rejection. The five-year follow-up showed that our multi-omics classification is a better prognostic factor than BOR. Therefore, the analysis reported here can serve as a guide for improved patient stratification and selection strategies in combination therapies involving ICI and immuno-modulatory agents, including DHA.

Longitudinal Whole Exome Sequencing^42^ and transcriptomic analysis contributed to shed light on molecular factors impacting on clinical response and on long term outcome after guadecitabine plus ipilimumab. At the mutational profile level, strong consistency across biopsies obtained at three time points in a 12-week time frame allowed us to identify significant associations of somatic mutations with response/resistance even with the small number of patients enrolled in the NIBIT-M4 trial.

Mutations in genes belonging to the Epithelial to Mesenchymal Transition (EMT) were enriched in NR patients. EMT is associated with a less favorable outcome in patients with cancer ^23^; it stimulates angiogenesis and is a tumor-intrinsic mechanism enhancing immunosuppression^43,44^. Recent studies reporting the genomics and transcriptomics features associated with ICI in melanoma have shown higher expression of several EMT genes in non-responder subjects^45^. Our analysis indicates that genomic alterations can drive these differences. We also observed that two NR patients harbored mutations in the gene *DNMT1*, which is the direct target of guadecitabine. One such mutation is a truncating event giving rise to the hypothesis that the loss of function of *DNMT1* hampers the immuno-modulatory effect of guadecitabine. Another defect in chromatin organization, gene *SETD2*, was observed in the NR group of treated patients. These two novel findings represent a first selection strategy to increase the success of similar studies^16,17^.

Interrogating the longitudinal gene expression data was also useful in evaluating how the immune context evolves during therapy. The differential activity of these pathways tends to increase during therapy, suggesting that immune surveillance promoted by ICI represses cell cycle genes together with differentiation pathways. One of the most interesting messages of our data is that the most evident difference between patients is the dynamic increase of the level of NK-cells and CD8 T-cells in patients that respond to therapy, rather than the tumor microenvironment composition at the baseline. When transcription-based signatures for response to ICI prediction such as TIDE^27^, IMPRES^25^, ICR^20^, and MIRACLE^9^ were interrogated, the only scores that significantly differentiated R and NR patients were ICR and MIRACLE signatures. However, this difference was not significant when just the baseline lesions were used. This suggests that additional factors contribute to clinical response, beyond the process of development of adaptive immunity, captured by these signatures. We reasoned that an immune-based stratification approach taking into account: a) the evidence of expression of genes associated with immunological constant of rejection (ICR) and; b) the amount of immuno-editing measured as the ratio of observed versus expected neo-antigens (GIE) could improve our ability to understand response and resistance to treatment. The presence of an adaptive immune response within tumors is accounted by the ICR^19,20^, a signature that predicts survival and response to ICI therapy in different tumors such as breast^21^, bladder, stomach, head and neck^20^, sarcoma^22^ and melanoma^9^. The importance of immuno-editing, and its association with survival and resistance has been extensively demonstrated in human primary tumors^38^ and in immune selection pressure on metastatic evolution^18^. The GIE score is a measure of the extent of immunoediting occurring in a tumor and it is obtained through comparison of “predicted neoantigens” (i.e., genes encoding putative HLA-class I-binding neoantigenic peptides, identified through integrative analysis of WES data and of HLA genotype of the patient) with “observed neoantigens” (i.e. actual expression of genes encoding neoantigens as obtained through transcriptomic data analysis). Patients whose tumors have a GIE score <1 indicate previous immunoediting (i.e., they have a number of expressed/observed neoantigens lower than the number of predicted neoantigens). The observation that the GIE score and the ICR were uncorrelated suggested that they are capturing complementary, yet distinct, attributes of anti-tumoral immunity. Therefore, their combination could be an effective means to achieve a more accurate quantification of effective cancer immune surveillance and of response to immunotherapy. Indeed, the ICR/GIE classification could stratify NIBIT-M4 patients into four subsets, and those with high ICR scores (>0) and a low (<1) GIE score showed the longest OS. In other words, the coupling of adaptive immunity with effective immunoediting is a relevant requisite for achieving long term clinical benefit from treatment. The prognostic significance of the ICR/GIE index was confirmed by the analysis of independent datasets from ICI-treated patients, suggesting that this is a robust classifier that captures crucial immunological processes acting in the context of different immunotherapy regimens. The activation of T cell-mediated immunity is dependent upon the recognition of tumor antigens on major histocompatibility complexes (MHC) of antigen-presenting cells^46^. Tumor antigen presentation by MHC class I is mediated by the coordinated expression of multiple genes. The differences between the High-ICR/GIE and High-ICR/Non-GIE, observed in our cohort and other independent cohorts, confirm that even in the presence of an adaptive immune response, tumor cells that develop defects in antigen processing or presentation can escape immune surveillance^37,42^.

The low number of analyzed cases, even with longitudinal multi-omics profiling, represents a limitation in our study, though its results help to guide further development of DHA/ICI combinations in solid tumors. Furthermore, the application of the ICR/GIE to multiple contexts at a pan-cancer level is needed to further validate the clinical relevance of our approach and findings. Collectively, though limited by the still scarce number of completed trials and enrolled patients, the available clinical results suggest and support further development of combinatorial approaches of DHA with ICI in cancer therapy in randomized clinical trials with extensive translational endpoints.

## Materials and Methods

### Study design, patient population, procedures, and outcomes

We conducted a milestone, 5-year follow-up analysis of patients enrolled in the NIBIT-M4 study; the study design, patient eligibility criteria, and treatment regimen have already been described (REF 12?). Briefly, the phase Ib, dose-escalation, single-center NIBIT-M4 study, enrolled pre-treated or untreated patients with unresectable Stage III or IV melanoma, to receive guadecitabine 30, 45, or 60 mg/m^2^/day s.c. on days 1-5 at week 0, 3, 6, 9, and ipilimumab 3 mg/kg i.v. on day 1 at week 1, 4, 7, 10, for 4 cycles. For this follow-up analysis, median OS, PFS, 5-year OS and PFS rate, and median DoR were assessed. Patients were classified as R or NR based on Disease Control. Tumor biopsies for correlative analyses were performed at baseline and at week 4 and week 12 on-treatment.

### Data collection, Library preparation, and sequencing

Isolation of total DNA/RNA and library preparation for RNA Sequencing and Reduced Representation Bisulfite Sequencing (RRBS) were performed as previously described^13^ at different time points of treatments (week0, week4, week12) for N=14 patients, including eight additional patients not available in the previous study. For Whole exome sequencing (WES), Nextera Flex for Enrichment solution (Illumina, San Diego, CA) in combination with SureSelect Human All Exon V7 probes (Agilent, Santa Clara, CA) was used for library preparation and generated libraries were sequenced on NovaSeq 6000 (Illumina, San Diego, CA) in 150 pair-end mode for biopsies of patients from 1 to 8; TruSeq Rapid Exome (Illumina, San Diego, CA, USA) was used for library preparation and generated libraries were sequenced on HiSeq 3000/4000 (Illumina, San Diego, CA) in 150 pair-end mode for biopsies of patients from 9 to 14.

### Data processing

#### Whole Exome Sequencing

Quality control of Whole exome sequencing (WES) was performed on raw data using fastQC (v. 0.11.8) (https://www.bioinformatics.babraham.ac.uk/projects/fastqc/).

Sequencing reads were aligned to Human reference genome (UCSC genome assembly GRCh37/hg19) using Burrows-Wheeler Aligner^47^, and then processed by GATK3^48^ for discarding low mapping quality reads and performing indel realignment.

Somatic single-nucleotide variants (SNVs) and indels calling were performed using Sentieon Genomic Tool 201911^49^. A virtual normal panel from 1000 Genomes Project^50^ was used to call SNVs and indels for tumor samples without a matched normal sample.

Putative false positive calls have been removed considering as filters: i) the variant-supporting read count greater than 2; ii) variant allele frequency greater than 0.05; iii) average variant position in variant-supporting reads (relative to read length) greater than 0.1 and lower than 0.9; iv) average distance to effective 3′ end of variant position in variant-supporting reads (relative to read length) greater than 0.2; v) fraction of variantsupporting reads from each strand greater than 0.01; vi) average mismatch quality difference (variant − reference) lower than 50; vii) average mapping quality difference (reference − variant) lower than 50. Annotation of SNVs and indels was performed using AnnoVar^51^ and SnpEff^52^. The functional effect of missense SNVs and in-frame indels was computed using Polyphen2 ^53^, SIFT ^54^ and PROVEAN ^55^ algorithms and variants predicted as damaging at least two of them were classified as pathogenic mutations. Somatic copy number was estimated from WES reads by CNVkit^56^ and GISTIC^57^ was applied for identifying genomic regions recurrently amplified or deleted. The nonsynonymous tumor mutational burden (TMB) was computed as the number of non-synonymous somatic mutations (single nucleotide variants and small insertions/deletions) per megabase in coding regions.

#### RNA Sequencing

Fastq quality was assessed using fastQC (v. 0.11.8) (https://www.bioinformatics.babraham.ac.uk/projects/fastqc/) and low quality reads were discarded. Sequence reads were aligned to Human reference genome (UCSC genome assembly GRCh38/hg38) using STAR (v. 2.7.0b)^58^, and the expression was quantified at gene level using featureCounts (v. 1.6.3), a countbased estimation algorithm^59^. Downstream analysis was performed in the R statistical environment as described below. Raw data were normalized according to sample-specific GC-content differences as described in EDAseq R package (v. 2.22.0)^60^. Differential expression analysis was performed using EdgeR R package (v. 3.30.3)^61^. Genes sorted according to log2 fold-change (log2FC) were used for performing Gene Set Enrichment Analysis (GSEA) of Gene Ontology (GO) Biological Processes (BP)^62^, as implemented in the clusterProfiler R package (v. 3.3.6)^63^.

#### RRB Sequencing

Reduced Representation Bisulfite Sequencing (RRBS) raw reads were trimmed for adaptor sequences using trim galore (v. 0.6.5) (http://www.bioinformatics.babraham.ac.uk/projects/trim_galore/) and filtered for low-quality sequences using fastQC (v. 0.11.8) (https://www.bioinformatics.babraham.ac.uk/projects/fastqc/). High quality trimmed reads were mapped to the Human reference genome (UCSC genome assembly GRCh38/hg38) using Bismark (v. 0.22.3) ^64^ with default parameters. Methylation data as β values were retrieved from bismark coverage outputs using R package RnBeads 2.0 (v. 2.6.0)^65^ with default parameters. Then, human GRCh38 annotated genes and promoters exhibiting differential DNA methylation between predefined groups of patient samples were identified using R package limma (v. 3.44.3)^66^.

### Prediction of immune response and tumor microenvironment deconvolution

ICR scores were computed for each sample using a single sample GSEA (ssGSEA) based on the Mann– Whitney–Wilcoxon Gene Set Test (MWW-GST)^67^ and the ICR signature *(IFNG, IRF1, STAT1,IL12B, TBX21, CD8A, CD8B, CXCL9, CXCL10, CCL5, GZMB, GNLY, PRF1, GZMH, GZMA, CD274/PD-L1, PDCD1, CTLA4, FOXP3*, and *IDO1)*^*24*^.

MIRACLE scores were computed using the MIRACLE R package as described in Turan et al. (2021)^9^. TIDE score was computed using TIDE command-line interface (https://github.com/jingxinfu/TIDEpy)^26,27^. IMPRES score was computed using *calc_impres* R function (https://github.com/Benjamin-Vincent-Lab/binfotron/)^68^. Estimation of immune and stromal subpopulation abundances was computed using MCP-counter^36^.

### TCR repertoire analysis from RNA Sequencing data

The docker implementation of MiXCR software (v. 3.0.13)^69^ was used to retrieve the VDJ repertoire from RNA Sequencing data. For the T cell receptor Beta locus (TRB), the clonality was calculated as

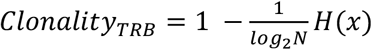

where *H*(*x*) is the Entropy computed as standard Shannon entropy as follow:

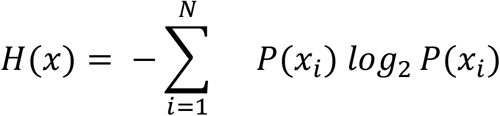

For a productive (in-frame) sequence *x*_*i*_, *P*(*x*_*i*_), is the ratio between the sequence count and total productive count and *N* is the number of productive unique in-frame sequences.

### Integrative analysis of RNASeq and RRBS data

Integration of gene expression and methylation data was performed as follows, according to the type of region level methylation data.

1. *Gene level methylation*. For each matched RNAseq-RRBS profiling sample, gene expression and methylation ranks were aggregated using robust rank aggregation (RRA) method implemented in the R package RobustRankAggreg (v. 1.2)^70^. A Single sample GSEA (ssGSEA) was computed using the Mann–Whitney–Wilcoxon Gene Set Test (MWW-GST) as previously described in Frattini et al., 2018 ^67^, starting from the expression-methylation aggregated rank and a manually curated collection of signatures downloaded from MSigDB ^71^, including GO BP, REACTOME-, BIOCARTA- and HALLMARK-pathways. Molecular pathway changes that occurred at different treatment time points in R and NR groups and were simultaneously mediated by expression and methylation were summarized as heatmap using R package complexHeatmap (v. 2.11.1)^72^. For visualization purposes, the resulting heatmap was obtained using the scaled Normalized Enrichment Score (NES) averaged in each time-response subgroup of the top differentially enriched and most statistically significant (FDR <0.01) signatures in R vs. NR patient comparison.
2. *Promoter level methylation*. R package SMITE (v. 1.16.0)^73^ was applied to identify functionally related genes with altered DNA methylation on promoters. Briefly, each differentially expressed gene (R vs. NR patients at each treatment time point week0, week4 and week12) previously computed using the EdgeR R package (v. 3.30.3)^61^ was associated with a promoter region [TSS − 1 kb, TSS + 500 bp] using UCSC GRCh38/hg38 refSeq transcripts coordinates. Each promoter was then associated with a set of overlapping regions from the differential methylation analysis previously computed on the same comparison using R package limma (v. 3.44.3)^66^. To identify genes whose expression is inversely correlated with promoter methylation, a score based on a weighted significance value (0.5 for expression and 0.3 for promoter methylation) was computed. Hypomethylated/up-regulated and hypermethylated/down-regulated genes for each comparison were then visualized as scatterplot using R package ggplot2 (v. 3.3.6). These modules of expression/methylation concordat genes were functionally analyzed through a GO BP pathway enrichment analysis within the R package SMITE. Significant categories (*p*-value < 0.05) were visualized as barplot using R package ggplot2.

### HLA typing and neoantigen prediction

HLA typing was performed from WES data using the docker implementation of Polysolver (v. 4)^74^. The neoantigen prediction tool pVACseq from pVACtools^75^ was run using the following predictors: MHCnuggetsI, NNalign, NetMHC, SMM, SMMPMBEC, and SMMalign.

Mutant-specific binders, relevant to the restricted HLA-I allele, are referred to as neoantigens, as previously described^76^. To infer neoantigens with high confidence, we considered only the mutated epitopes with a median IC_50_ binding affinity across all prediction algorithms used < 500 nM, with a corresponding wildtype epitope with a median IC_50_ binding affinity > 500 nM.

### Genetic ImmunoEditing (GIE) score

The Genetic ImmunoEditing (GIE) score was computed by taking the ratio between the number of observed (O) in a patient versus the number of expected (E) for that patient:

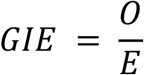

The observed number of neoantigens *O* was obtained from the output of pVACtools^75^, filtered according to the criteria described above.

The expected number of neoantigens *E* was computed as a function of the number of nonsynonymous mutations by fitting a linear regression model using the *lm* function of R package stats (v. 4.0.2) trained with the data of our cohort, using the number of neoantigens as dependent variable. We assumed that samples that show a frequency of neoantigens lower than expected (i.e., lower GIE values) have evidence of immunoediting.

Based on GIE and ICR scores, we classified samples as “High-ICR/GIE” (ICR > 0 and GIE < 1), “HighICR/Non-GIE” (ICR > 0 and GIE > 1), “Low-ICR/GIE” (ICR < 0 and GIE < 1), “Low-ICR/Non-GIE” (ICR < 0 and GIE > 1).

### Survival analysis

Survival curves were estimated using the survival R package (v. 3.2-10) and plotted using the Kaplan-Meier method, implemented in the survminer (v. 0.4.9) R package. Log-rank tests were used to compare curves between groups using.

### IHC analysis

Serial 3 μm formalin-fixed paraffin-embedded tissue sections were stained using an AutostainerPlus (Dako). Antigen retrieval and deparaffinization were carried out on a PT-Link (Dako) using the EnVision FLEX Target Retrieval Solutions (Dako). Endogenous peroxidase and non-specific staining were blocked with H202 3% (Gifrer, 10603051) and Protein Block (Dako, X0909) respectively. The antibodies used are: anti-CD8 clone C8/144B (M7103, Dako) and anti-HLA Class I, clone EMR8-5 (ab70328, abcam). The HRP labeled polymer conjugated EnVision+ Single Reagent (Dako, K4001) was used as a secondary antibody. Peroxidase activity was detected using 3-amino-9-ethylcarbazole substrate (Vector Laboratories, SK-4200). All stained slides were digitalized with a NanoZoomer scanner (Hamamatsu).

### NanoString

Expression of genes belonging to several immune-related signatures was assessed by a custom-designed NanoString nCounter multiplex CodeSet enabling determination of 364 genes. The gene signatures were selected for providing information on B cell content and differentiation, TLS formation, follicular T helper cells, TEX subsets, tumor-associated endothelial cells, ICB response and guadecitabine-specific gene upregulation. For NanoString experiments, panel probes (capture and report) and 200 ng of RNA were hybridized overnight at 65 °C for 16 h. Samples were scanned at maximum scan resolution capabilities (555 FOV) using the nCounter Digital Analyzer. Quality control of samples, data normalization and data analysis were performed using nSolver software 4.0 (NanoString Technologies).

### ICR/GIE validation in independent cohorts

Molecular and clinical data for three independent immunogenomic datasets of melanoma patients treated with ICI^4,39,40^ were obtained from cBioportal^77^. A total of 83 patients for which all required data were available (gene expression, mutation/neoantigen loads, treatment response and overall survival) were selected and grouped in responder (CR: complete response, PR: partial response, SD: stable disease, LB: long-term benefits) and non-responder (PD: progression disease, NB: minimal or no-benefits) according to the treatment outcome as described in the corresponding original studies. Integrated gene expression matrix was batch corrected using the *removeBatchEffect* function implemented in R package limma (v. 3.44.3)^66^. ICR and Genetic Immunoediting scores were computed as previously described. Differential expression analysis was performed between “High ICR/GIE” and “High ICR/Non-GIE” classes using a wilcoxon test. Genes sorted according to log2 fold-change (log2FC) were used for performing Gene Set Enrichment Analysis (GSEA) of Gene Ontology (GO) Biological Processes (BP)^62^, as implemented in the clusterProfiler R package (v. 3.3.6)^63^. Selected enriched GO:BP categories (FDR < 0.01) were visualized as a barplot. Survival analysis was performed as previously described.

## Data Availability

The processed NanoString dare are available via the GEO accession number: GSE211645 [https://www.ncbi.nlm.nih.gov/geo/query/acc.cgi?acc=GSE211645; token mjghgimevhmpbkx]. Raw data of Whole Exome Sequencing, RNA-Sequencing and Reduced Representation Bisulfite Sequencing are available on the European Genome-phenome Archive under accession number EGAS00001006736.

https://ega-archive.org/studies/EGAS00001006736

## Supplementary Information

### Figures

**Figure S1**. *NIBIT-M4 cohort*.

Overview of sample profiling and clinical information for N=14 patients of NIBIT-M4 trial.

**Figure S2**.

ICI response prediction scores between responder and non-responder samples at baseline (*p*-value of Student’s t-Test) (A). Heatmap of expression z-scores (from NANOSTRING gene expression assay) computed for selected pathways for all patient samples. ICR classification in “High ICR” (ICR enrichment NES > 0) and “Low ICR” (ICR enrichment NES < 0) is reported (B). Volcano plot of differentially expressed genes between responders and non-responders at baseline (week0), four (week4) and twelve (week12) weeks after treatments. Guadecitabine target genes are labeled (C).

**Figure S3**.

Boxplot of GIE score at baseline (left) and all time points (right) between responder and non-responder samples (*p*-value of Student’s t-Test) (A). Scatterplot of ICR score by for responder and non-responder samples at baseline (left) and their proportion (right) after classification as ICR/GIE classes in responder and non-responder groups (*p*-value from Pearson’s chi-squared test statistic) (B).

**Figure S4**.

Barplot of frequencies of HLA-I score (calculated by multiplying the score for staining intensity with the percentage of positive cells from IHC staining and then grouped into negative (=0), low (< median) and high (>=median)) for samples classified as High ICR/GIE or High ICR/Non-GIE at week12 (*p*-value from Pearson’s chi-squared test statistic) (A). Kaplan Meier for PFS by patients classified as High ICR/GIE or High ICR/Non-GIE at week12 (top-left) and responder or non-responder (top-right). Kaplan Meier for OS by patients classified according to ICR/GIE subtypes (bottom-left) and for PFS by patients classified as responder or non-responder (bottom-right). Time is indicated in months and censor points are indicated by vertical lines (B).

**Figure S5**. Characterization of baseline (Week 0), Week 4 and Week 12 tumor biopsies for ICR and GIE scores, for CD8+ density in the tumor core and for level of expression of HLA Class I by immunohistochemistry (A). Localization and evolution of neoplastic lesions in patient #11 (B)

### Tables

**Table S1**. Somatic mutations of NIBIT-M4 cohort.

**Table S2**. Differentially expressed genes between R vs. NR at baseline, week4 and week12 after treatment **Table S3**. Gene Set Enrichment analysis results from R vs. NR comparison at baseline, week4 and week12 after treatment

**Table S4**. Predictive scores of response to ICI for NIBIT-M4 samples

**Table S5**. GIE scores for NIBIT-M4 samples

## Ethics Statement

The study was conducted in accordance with the ethical principles of the Declaration of Helsinki and the International Conference on Harmonization of Good Clinical Practice. The protocol was approved by the independent ethics committee of the University Hospital of Siena (Siena, Italy). All participating patients (or their legal representatives) provided signed-informed consent before enrolment. The trial was registered with European Union Drug Regulating Authorities ClinicalTrials EudraCT, number 2015-001329-17, and with ClinicalTrials.gov, number NCT02608437.

## Acknowledgements

The research leading to these results has received funding from AIRC under 5 per Mille 2018 - ID.21073 project – P.I. Maio Michele, G.L. Anichini Andrea, G.L. Ceccarelli Michele. The research leading to these results has received funding from AIRC under IG 2018 - ID. 21846 project – P.I. Ceccarelli Michele. This work was also supported by the Italian Ministry of Research Grant PRIN 2017XJ38A4_004. The authors wish to tank the EPigenetic Immune-oncology Consortium AIRC (EPICA) investigators (Giuseppe Palmieri, Daniela Massi and Ulrich Pfeffer) for helpful discussion of data and critical review of the manuscript.

## Author contributions

Conceptualization: M.T.R. N., A.M.D.G., A.A., M.M. and M.C.; Methodology: M.T.R. N., A.M.D.G., W.H.F., C. S.-F., R.M., M.F. L., D.B., A.A., M.M. and M.C.; Investigation: M.T.R. N., A.M.D.G., F.P.C., A.C., S.C., W.H.F., C. S.-F., S.B., G.P., E.S., M.M. and M.C.; Analysis: M.T.R. N., A.M.D.G., F.P.C., A.C., G.S., M.C.C., S.C., W.H.F., C. S.-F., S.B., G.P., M.M. and M.C.; Writing: M.T.R. N., A.M.D.G., R.M., A.A., M.M. and M.C; Supervision: M.M. and M.C.

