## supplementary Figures for "Guadecitabine plus ipilimumab in unresectable melanoma: five-year follow-up and correlation with integrated, multiomic analysis in the NIBIT-M4 trial"

| Patient | Sex | Dose | BOR | WES | | | | RNASeq | | | RRBS | | | NanoString | | |
|---|---|---|---|---|---|---|---|---|---|---|---|---|---|---|---|---|
| | | | | Normal | W0 | W4 | W12 | W0 | W4 | W12 | W0 | W4 | W12 | W0 | W4 | W12 |
| 1 | F | 30 | NR | ✓ | ✓ | ✓ | ✓ | ✓ | ✓ | ✓ | ✓ | ✓ | ✓ | ✓ | ✓ | |
| 2 | M | 30 | NR | ✓ | ✓ | ✓ | ✓ | ✓ | ✓ | ✓ | ✓ | ✓ | ✓ | ✓ | ✓ | ✓ |
| 3 | M | 30 | R | ✓ | ✓ | ✓ | ✓ | ✓ | ✓ | ✓ | ✓ | ✓ | ✓ | ✓ | ✓ | ✓ |
| 4 | M | 45 | R | ✓ | ✓ | ✓ | ✓ | ✓ | ✓ | ✓ | ✓ | ✓ | ✓ | ✓ | ✓ | ✓ |
| 5 | M | 45 | R | ✓ | ✓ | ✓ | | ✓ | ✓ | | ✓ | ✓ | | ✓ | ✓ | |
| 6 | M | 45 | NR | ✓ | ✓ | ✓ | ✓ | ✓ | ✓ | ✓ | ✓ | ✓ | ✓ | ✓ | ✓ | ✓ |
| 7 | M | 45 | NR | ✓ | ✓ | ✓ | ✓ | ✓ | ✓ | ✓ | ✓ | ✓ | ✓ | ✓ | ✓ | ✓ |
| 8 | M | 45 | NR | ✓ | ✓ | ✓ | ✓ | ✓ | ✓ | ✓ | ✓ | ✓ | ✓ | ✓ | ✓ | ✓ |
| 9 | M | 45 | NR | | ✓ | ✓ | ✓ | ✓ | ✓ | ✓ | ✓ | ✓ | ✓ | ✓ | ✓ | ✓ |
| 10 | M | 60 | R | | ✓ | ✓ | ✓ | ✓ | ✓ | ✓ | ✓ | ✓ | ✓ | ✓ | ✓ | ✓ |
| 11 | F | 60 | R | | ✓ | ✓ | ✓ | ✓ | ✓ | ✓ | ✓ | ✓ | ✓ | ✓ | ✓ | ✓ |
| 12 | M | 60 | NR | | ✓ | ✓ | ✓ | ✓ | ✓ | ✓ | ✓ | ✓ | ✓ | ✓ | ✓ | ✓ |
| 13 | M | 60 | R | | ✓ | ✓ | ✓ | ✓ | ✓ | ✓ | ✓ | ✓ | ✓ | ✓ | ✓ | ✓ |
| 14 | M | 60 | NR | | ✓ | ✓ | ✓ | ✓ | ✓ | ✓ | ✓ | ✓ | ✓ | ✓ | ✓ | ✓ |

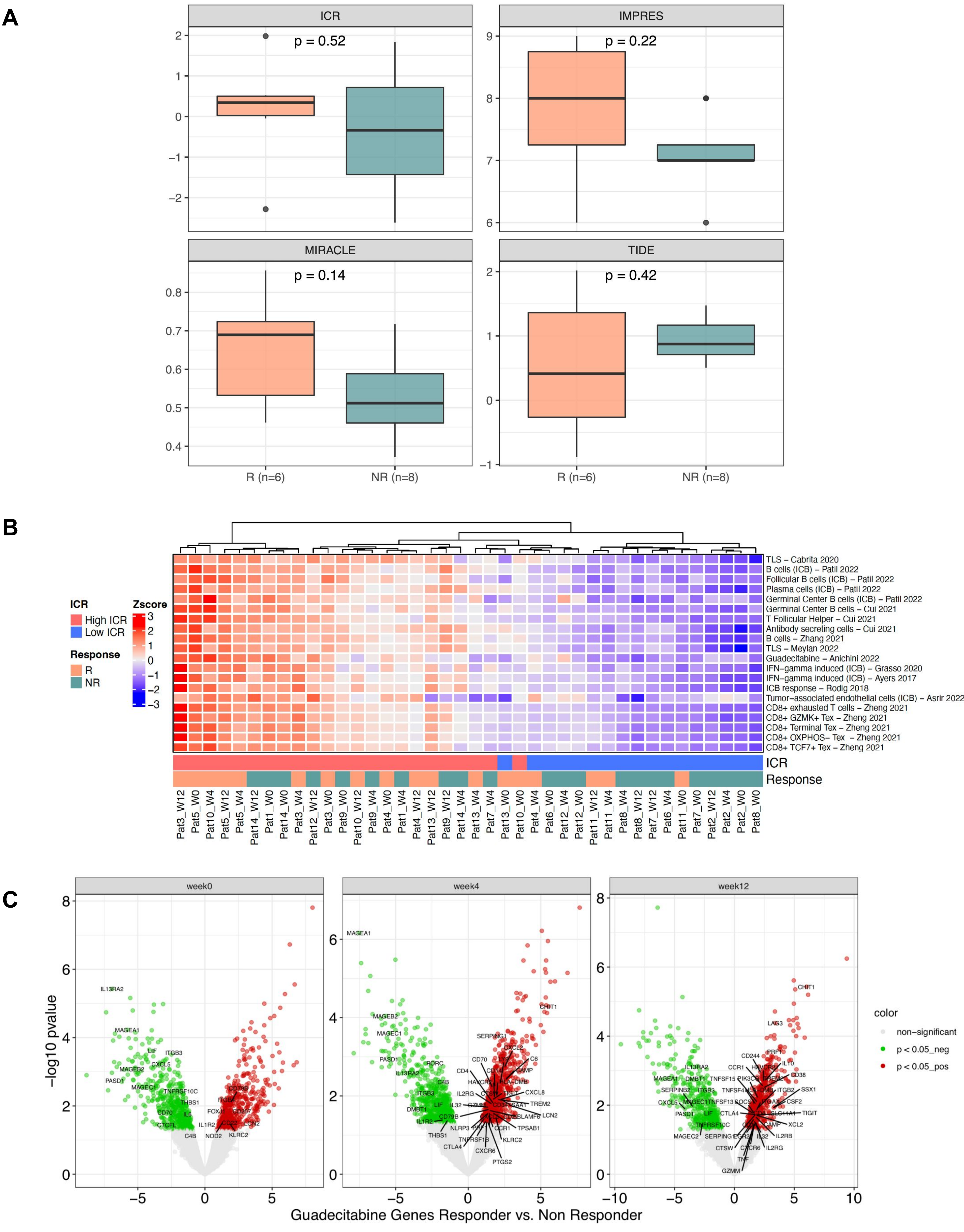
A
ICR
p = 0.52
IMPRES
p = 0.22
MIRACLE
p = 0.14
TIDE
p = 0.42
R (n=6)
NR (n=8)
R (n=6)
NR (n=8)
B
ICR
High ICR
Low ICR
Response
R
NR
Zscore
TLS – Cabrita 2020
B cells (ICB) – Patil 2022
Follicular B cells (ICB) – Patil 2022
Plasma cells (ICB) – Patil 2022
Germinal Center B cells (ICB) – Patil 2022
Germinal Center B cells – Cui 2021
T Follicular Helper – Cui 2021
Antibody secreting cells – Cui 2021
B cells – Zhang 2021
TLS – Meylan 2022
Guadecitabine – Anichini 2022
IFN–gamma induced (ICB) – Grasso 2020
IFN–gamma induced (ICB) – Ayers 2017
ICB response – Rodig 2018
Tumor–associated endothelial cells (ICB) – Asrir 2022
CD8+ exhausted T cells – Zheng 2021
CD8+ GZMK+ Tex – Zheng 2021
CD8+ Terminal Tex – Zheng 2021
CD8+ OXPHOS– Tex – Zheng 2021
CD8+ TCF7+ Tex – Zheng 2021
ICR
Response
Pat3_W12 Pat5_W0 Pat10_W4 Pat5_W12 Pat5_W4 Pat14_W12 Pat1_W0 Pat14_W0 Pat3_W4 Pat12_W12 Pat3_W0 Pat9_W0 Pat10_W12 Pat9_W4 Pat4_W0 Pat1_W4 Pat4_W12 Pat13_W12 Pat9_W12 Pat14_W4 Pat13_W4 Pat7_W4 Pat13_W0 Pat10_W0 Pat4_W4 Pat6_W0 Pat12_W4 Pat12_W0 Pat11_W12 Pat11_W4 Pat8_W4 Pat8_W12 Pat7_W12 Pat6_W4 Pat11_W0 Pat7_W0 Pat2_W12 Pat2_W4 Pat2_W0 Pat8_W0
C
week0
week4
week12
–log10 pvalue
Guadecitabine Genes Responder vs. Non Responder
color
non–significant
p < 0.05_neg
p < 0.05_pos

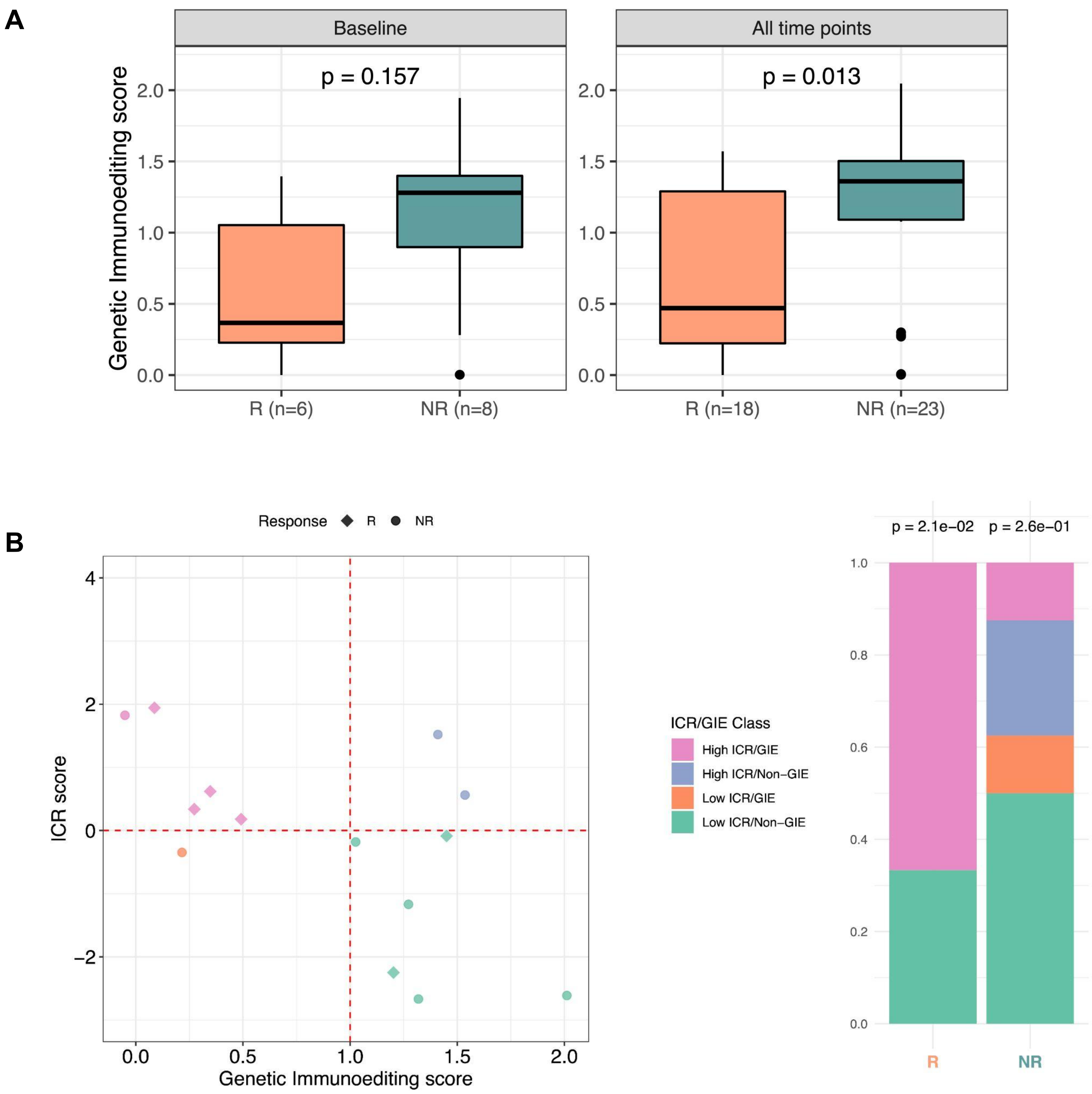
A
Baseline
All time points
p = 0.157
p = 0.013
Genetic Immunoediting score
2.0
1.5
1.0
0.5
0.0
R (n=6)
NR (n=8)
R (n=18)
NR (n=23)
B
Response
R
NR
ICR score
4
2
0
−2
0.0
0.5
1.0
1.5
2.0
Genetic Immunoediting score
ICR/GIE Class
High ICR/GIE
High ICR/Non−GIE
Low ICR/GIE
Low ICR/Non−GIE
p = 2.1e−02
p = 2.6e−01
1.0
0.8
0.6
0.4
0.2
0.0
R
NR

A

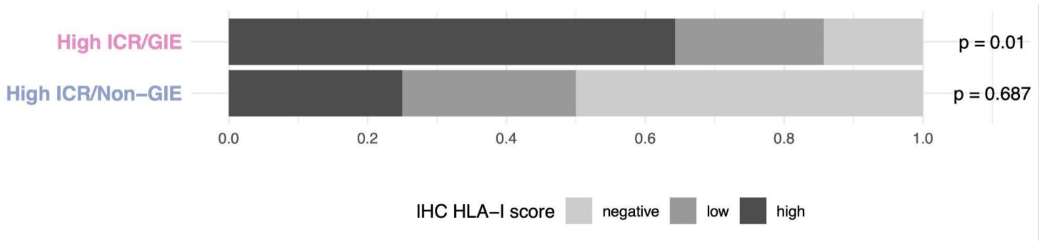
High ICR/GIE
High ICR/Non−GIE
p = 0.01
p = 0.687
0.0
0.2
0.4
0.6
0.8
1.0
IHC HLA−I score
negative
low
high

B

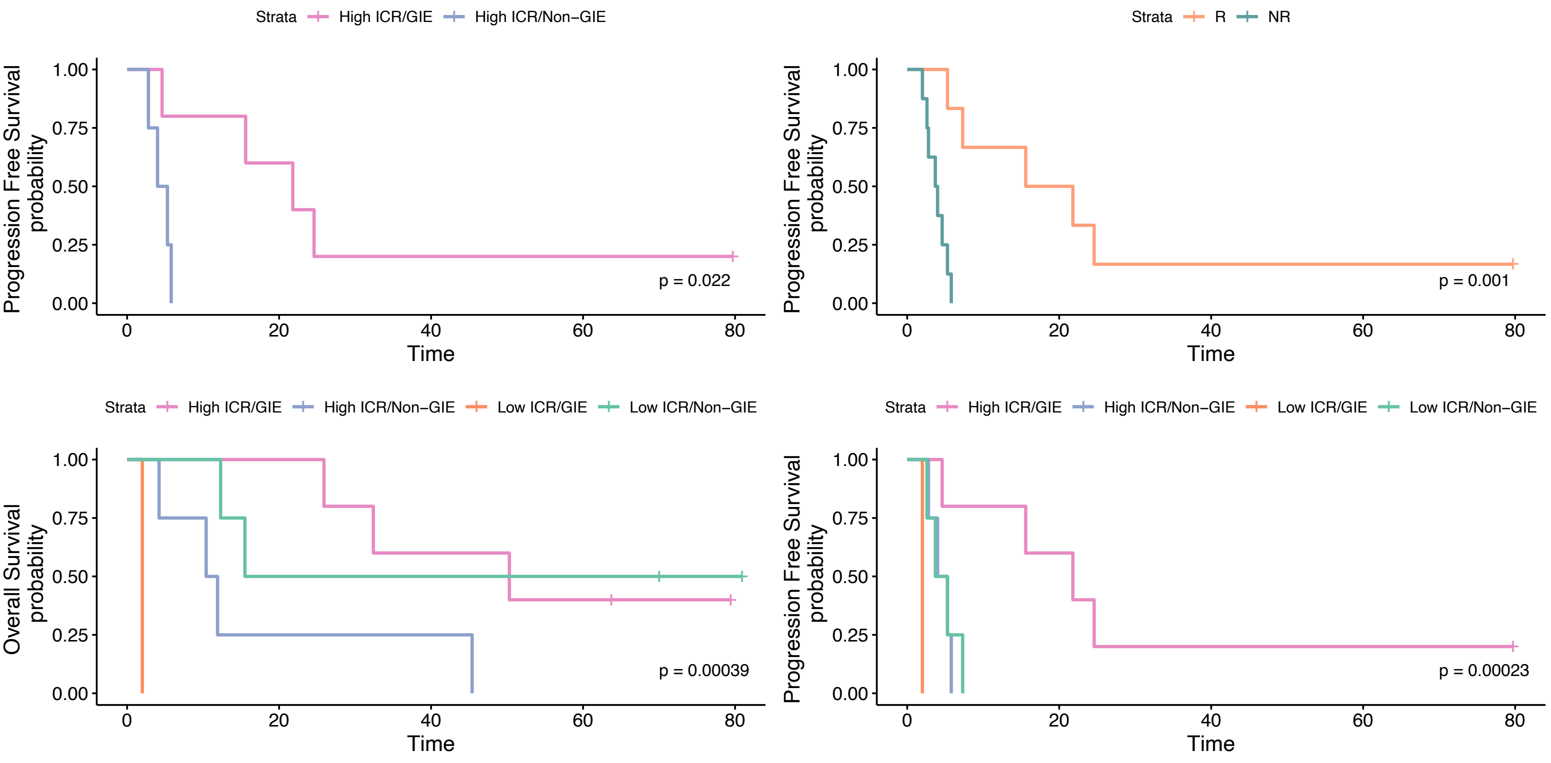
Strata
High ICR/GIE
High ICR/Non−GIE
Progression Free Survival probability
p = 0.022
Time
Strata
R
NR
Progression Free Survival probability
p = 0.001
Time
Strata
High ICR/GIE
High ICR/Non−GIE
Low ICR/GIE
Low ICR/Non−GIE
Overall Survival probability
p = 0.00039
Time
Strata
High ICR/GIE
High ICR/Non−GIE
Low ICR/GIE
Low ICR/Non−GIE
Progression Free Survival probability
p = 0.00023
Time

**A**

| Patient_ID | Timing of biopsy | Clinical response | ICR | GIE | CD8$^{+}$ density, tumor core (cells/mm$^{2}$) | Tumor HLA Class I Grade (0-3) | Agreement of clinical response with ICR/GIE/CD8/HLA profile | Possible interpretation |
|---|---|---|---|---|---|---|---|---|
| 3 | Week 0 | R | HIGH ICR | GIE | 1460 | 3 | YES | Response associated with High ICR/GIE classification, high level of CD8$^{+}$ infiltrate and retained tumor HLA class I expression |
| 3 | Week 4 | R | HIGH ICR | GIE | 639 | 3 | | |
| 3 | Week 12 | R | HIGH ICR | GIE | 1257 | 3 | | |
| 4 | Week 0 | R | HIGH ICR | GIE | 487 | 3 | YES | Response associated with High ICR/GIE classification, high level of CD8$^{+}$ infiltrate and retained tumor HLA class I expression |
| 4 | Week 4 | R | LOW ICR | NON-GIE | 106 | 3 | | |
| 4 | Week 12 | R | HIGH ICR | GIE | 912 | 3 | | |
| 5 | Week 0 | R | HIGH ICR | GIE | 1476 | 0 | YES | Response associated with High ICR/GIE classification, high level of CD8$^{+}$ infiltrate, in spite of defective HLA class I expression |
| 5 | Week 4 | R | HIGH ICR | GIE | 1139 | 2 | | |
| 5 | Week 12 | R | HIGH ICR | GIE | 1015 | 1 | | |
| 10 | Week 0 | R | HIGH ICR | GIE | 263 | 3 | YES | Response associated with High ICR/GIE classification, high level of CD8$^{+}$ infiltrate and retained tumor HLA class I expression |
| 10 | Week 4 | R | HIGH ICR | GIE | 1087 | 3 | | |
| 10 | Week 12 | R | HIGH ICR | GIE | 493 | 2 | | |
| 11 | Week 0 | R | LOW ICR | NON-GIE | 29 | 1 | NO | Response not associated with the observed ICR/GIE/CD8/HLA profile, but explained by clinical behaviour of tumor biopsies and target lesions in a patient with SD response (see panel B) |
| 11 | Week 4 | R | LOW ICR | NON-GIE | 82 | 2 | | |
| 11 | Week 12 | R | LOW ICR | NON-GIE | 680 | NA | | |
| 1 | Week 0 | NR | HIGH ICR | GIE | 564 | 1 | YES | Lack of response associated with low level of CD8$^{+}$ infiltrate and defective tumor HLA class I expression in spite of High ICR/GIE classification |
| 1 | Week 4 | NR | HIGH ICR | GIE | 153 | 0 | | |
| 1 | Week 12 | NR | HIGH ICR | GIE | 171 | 2 | | |
| 2 | Week 0 | NR | LOW ICR | NON-GIE | 65 | 0 | YES | Lack of response associated with Low ICR/Non GIE classification, low level of CD8$^{+}$ infiltrate and defective tumor HLA class I expression |
| 2 | Week 4 | NR | LOW ICR | NON-GIE | 142 | 1 | | |
| 2 | Week 12 | NR | LOW ICR | NON-GIE | 194 | 0 | | |
| 6 | Week 0 | NR | LOW ICR | GIE | 52 | 1 | YES | Lack of response associated with Low ICR/GIE classification, low level of CD8$^{+}$ infiltrate and defective tumor HLA class I expression |
| 6 | Week 4 | NR | LOW ICR | GIE | 86 | 2 | | |
| 7 | Week 0 | NR | LOW ICR | NON-GIE | 9 | 3 | YES | Lack of response associated with Low ICR/ Non-GIE classification, low level of CD8$^{+}$ infiltrate, in spite of retained tumor HLA class I expression |
| 7 | Week 4 | NR | HIGH ICR | NON-GIE | NA | 3 | | |
| 7 | Week 12 | NR | LOW ICR | NON-GIE | 107 | 3 | | |
| 8 | Week 0 | NR | LOW ICR | NON-GIE | 15 | 1 | YES | Lack of response associated with Low ICR/ Non-GIE classification, low level of CD8$^{+}$ infiltrate, in spite of retained tumor HLA class I expression |
| 8 | Week 4 | NR | LOW ICR | NON-GIE | 107 | 3 | | |
| 8 | Week 12 | NR | LOW ICR | NON-GIE | 201 | 3 | | |
| 9 | Week 0 | NR | HIGH ICR | NON-GIE | 550 | 1 | YES | Lack of response associated with High ICR/ Non-GIE classification, and loss of HLA class I on tumor, in spite of high level of CD8$^{+}$ infiltrate |
| 9 | Week 4 | NR | HIGH ICR | NON-GIE | 252 | 0 | | |
| 9 | Week 12 | NR | HIGH ICR | NON-GIE | 922 | 0 | | |

**B**

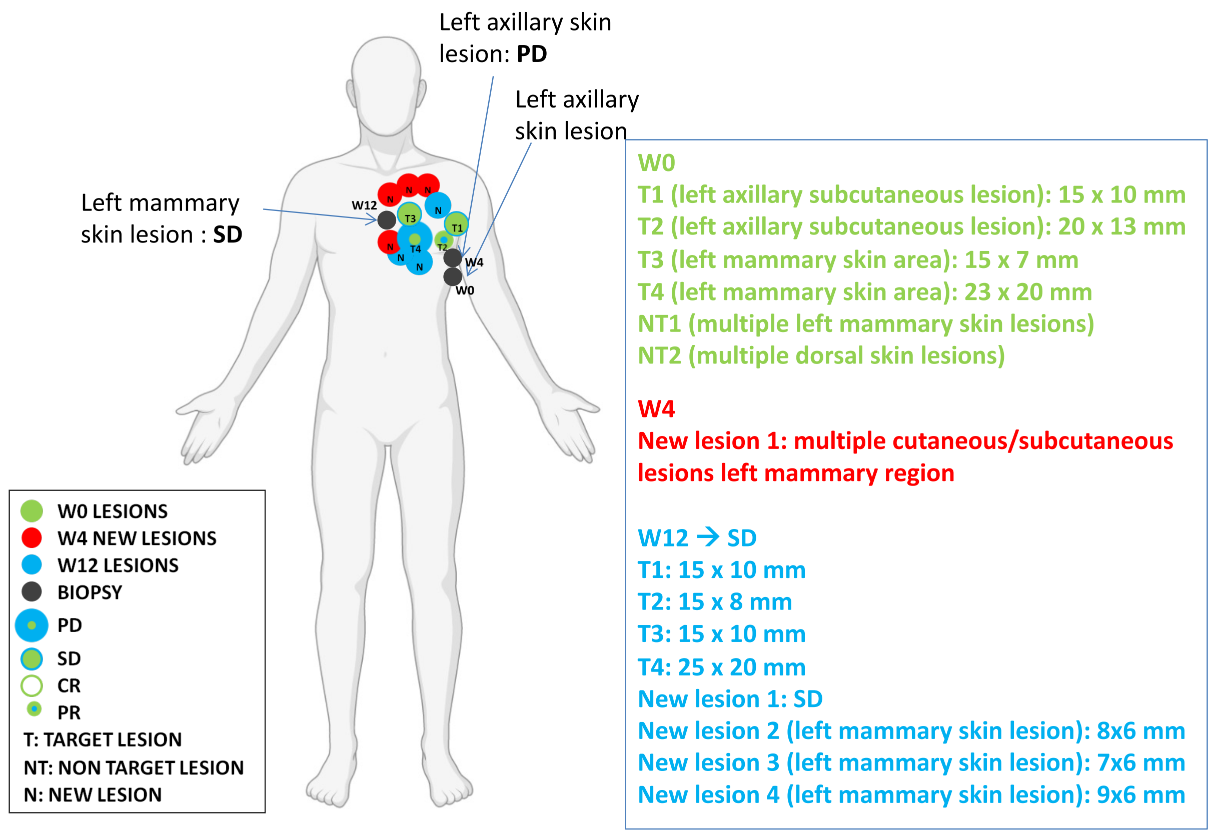
